# Predictive Models for Secondary Epilepsy in Patients with Acute Ischemic Stroke Within One Year

**DOI:** 10.1101/2024.06.27.24309564

**Authors:** Liu Jinxin, He Haoyue, Wang Yanglingxi, Du Jun, Liang Kaixin, Xue Jun, Liang Yidan, Chen Peng, Tian Shanshan, Deng Yongbing

## Abstract

**Objective:** Post-stroke epilepsy (PSE) is a major complication that worsens both prognosis and quality of life in patients with ischemic stroke. This study aims to develop an interpretable machine learning model to predict PSE using medical records from four hospitals in Chongqing.

**Methods:** We collected and analyzed medical records, imaging reports, and laboratory test results from 21,459 patients diagnosed with ischemic stroke. Traditional univariable and multivariable statistical analyses were performed to identify key predictive factors. The dataset was divided into a 70% training set and a 30% testing set. To address class imbalance, the Synthetic Minority Oversampling Technique combined with Edited Nearest Neighbors was used. Nine widely applied machine learning algorithms were evaluated and compared using relevant prediction metrics. SHAP (SHapley Additive exPlanations) was used to interpret the model, assessing the contributions of different features.

**Results:** Regression analyses showed that complications such as hydrocephalus, cerebral hernia, and deep vein thrombosis, as well as brain regions (frontal, parietal, and temporal lobes), significantly contributed to PSE. Factors like age, gender, NIH Stroke Scale (NIHSS) scores, and laboratory results such as WBC count and D-dimer levels were associated with a higher risk of PSE. Among the machine learning models, tree-based methods such as Random Forest, XGBoost, and LightGBM demonstrated strong predictive performance, achieving an AUC of 0.99.

**Conclusion:** Our model successfully predicts PSE risk, with tree-based models showing superior performance. The NIHSS score, WBC count, and D-dimer were identified as the most important predictors.

## Introduction

Stroke is the second leading cause of death globally, with an annual mortality of approximately 5.5 million, and it is also the leading cause of disability, accounting for 50% of cases worldwide [1]. Ischemic stroke comprises about 80% of all stroke cases [2][3]. Post-stroke epilepsy (PSE) is a common complication, with studies reporting that 3-30% of stroke patients develop epilepsy, which adversely affects their prognosis and quality of life [4]. PSE can worsen cognitive, psychiatric, and physical impairments already caused by cerebrovascular disease and related conditions [5]. The highest incidence of PSE occurs within the first year after an acute stroke, accounting for nearly half of the cases [2]. Thus, early prediction and intervention for PSE, especially in ischemic strokes, are critical.

Currently, most studies rely on clinical data to build statistical models using survival analysis, Cox regression [2][6], and multiple linear regression [7] to create basic models for PSE prediction. Last year, Lin et al. developed a radiomics-based model that outperformed conventional clinical models in predicting PSE related to intracerebral hemorrhage (ICH). They suggested that a combined radiomics-clinical model could improve the assessment of individual PSE risk after the first occurrence of ICH, facilitating early diagnosis and treatment [8]. However, subsequent research raised concerns about the use of radiomics, indicating a need for further investigation [9]. Overall, research on PSE prediction remains limited, with most studies focusing on specific risk factors [10][11][8][12] and building simple models, without proposing more comprehensive and scientifically robust prediction models.

Machine learning has gained attention as a powerful tool for building medical models due to its ability to process large datasets and complex information. It has been increasingly applied in neuroscience and clinical prediction [13][14][15]. Previous studies have used machine learning to explore post-stroke cognitive impairments [16], predict stroke and myocardial infarction risks in large artery vasculitis patients [14], develop post-stroke depression models based on liver function tests [17], and predict hematoma expansion in traumatic brain injury (TBI) [18].

Machine learning models can automatically manage both linear and complex nonlinear relationships between variables and offer insights into how different factors contribute to the prediction target—something that is difficult for traditional statistical models. However, machine learning requires substantial amounts of data and is prone to overfitting with small sample sizes. The quality and volume of input data are critical for the algorithm to detect underlying patterns and make accurate predictions.

This study aims to identify key risk factors from various features extracted from the clinical records and test data of ischemic stroke patients. Using these features, we will develop a machine learning-based prediction model for PSE. By leveraging early admission data, we seek to automatically predict the likelihood of PSE occurrence and provide guidance for clinical decision-making and patient care.

## Result

### Filling of missing data

Missing values were filled using a Random Forest (RF) model, handling one feature at a time. The imputed features were: Plt, WBC, RBC, HbA1c, CRP, TG, LDL, HDL, AST, ALT, bilirubin, albumin, urea, creatinine, BUA, PT, APTT, TT, INR, D-dimer, fibrinogen, CK, CK-MB, LDH, HBDH, IMA, lactate, anion gap, TCO2, and NIHSS.

### Characteristics of study participants

A total of 21,459 patients were included in the study. The training set consisted of 15,021 patients, with a PSE incidence of 4.3%. The test set contained 6,438 patients, also with a 4.3% incidence of PSE. The external validation cohort included 536 patients from three hospitals. The statistical details of the clinical characteristics are presented in Table 1.

**Table 1.**
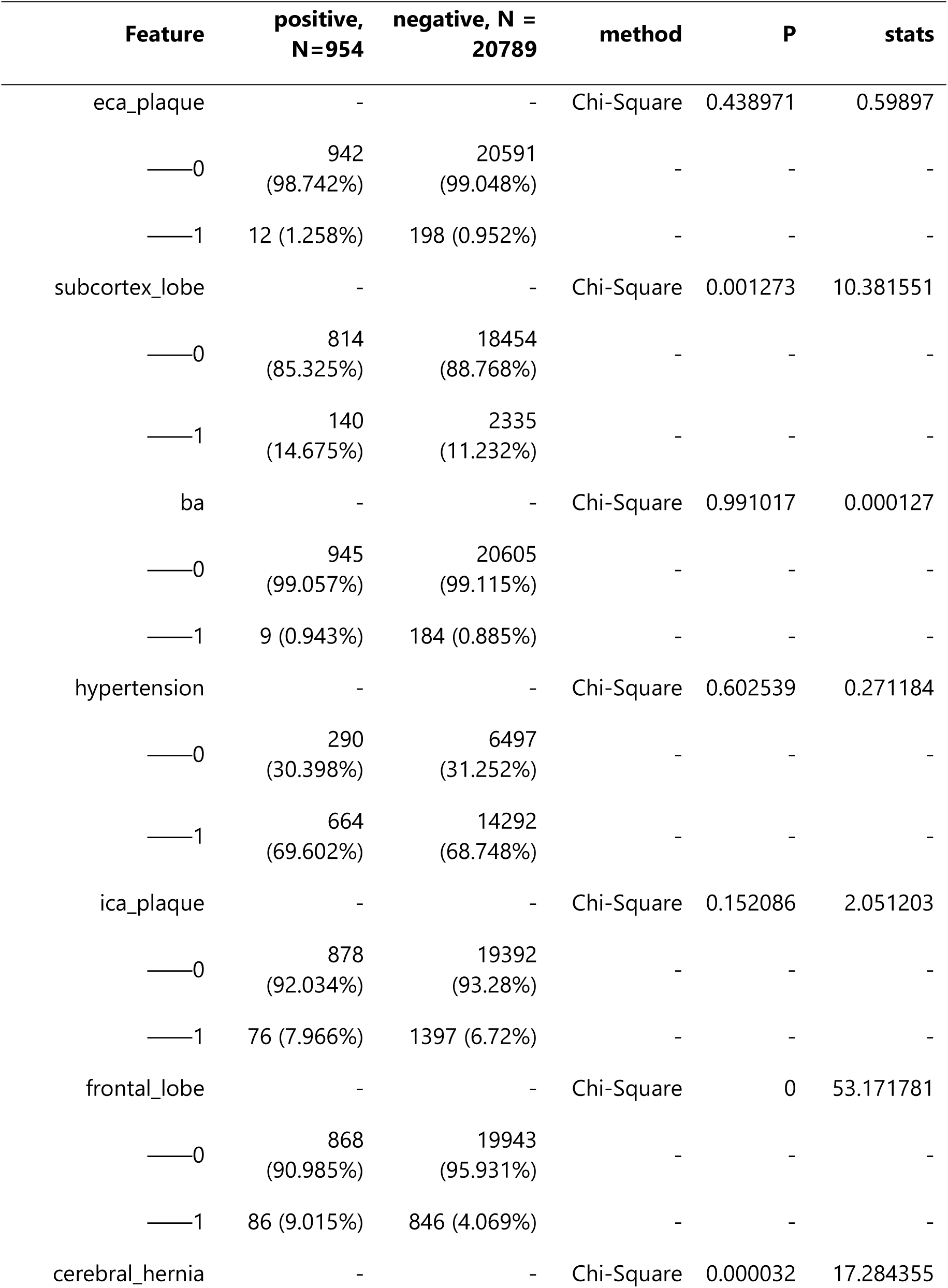
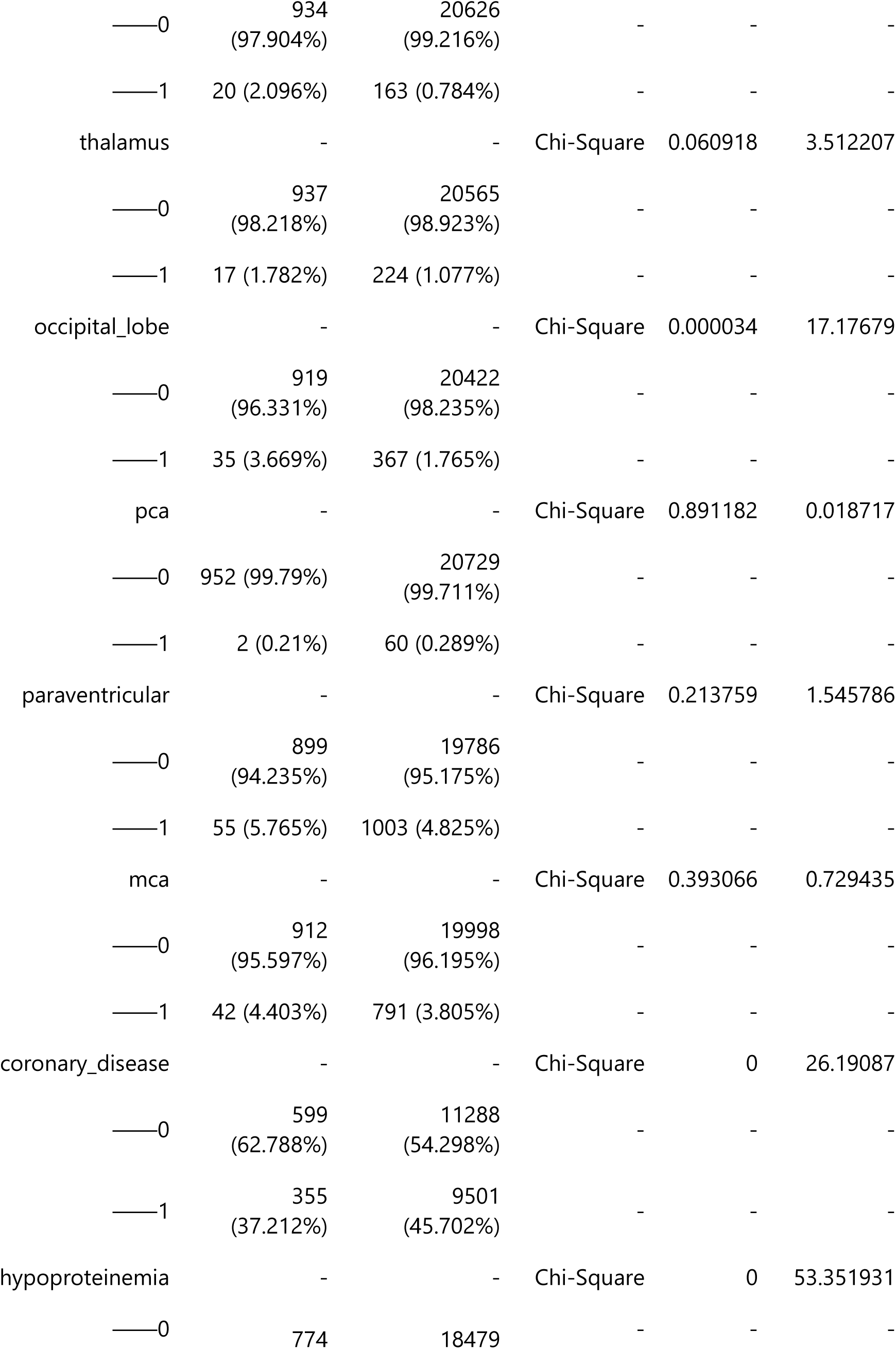
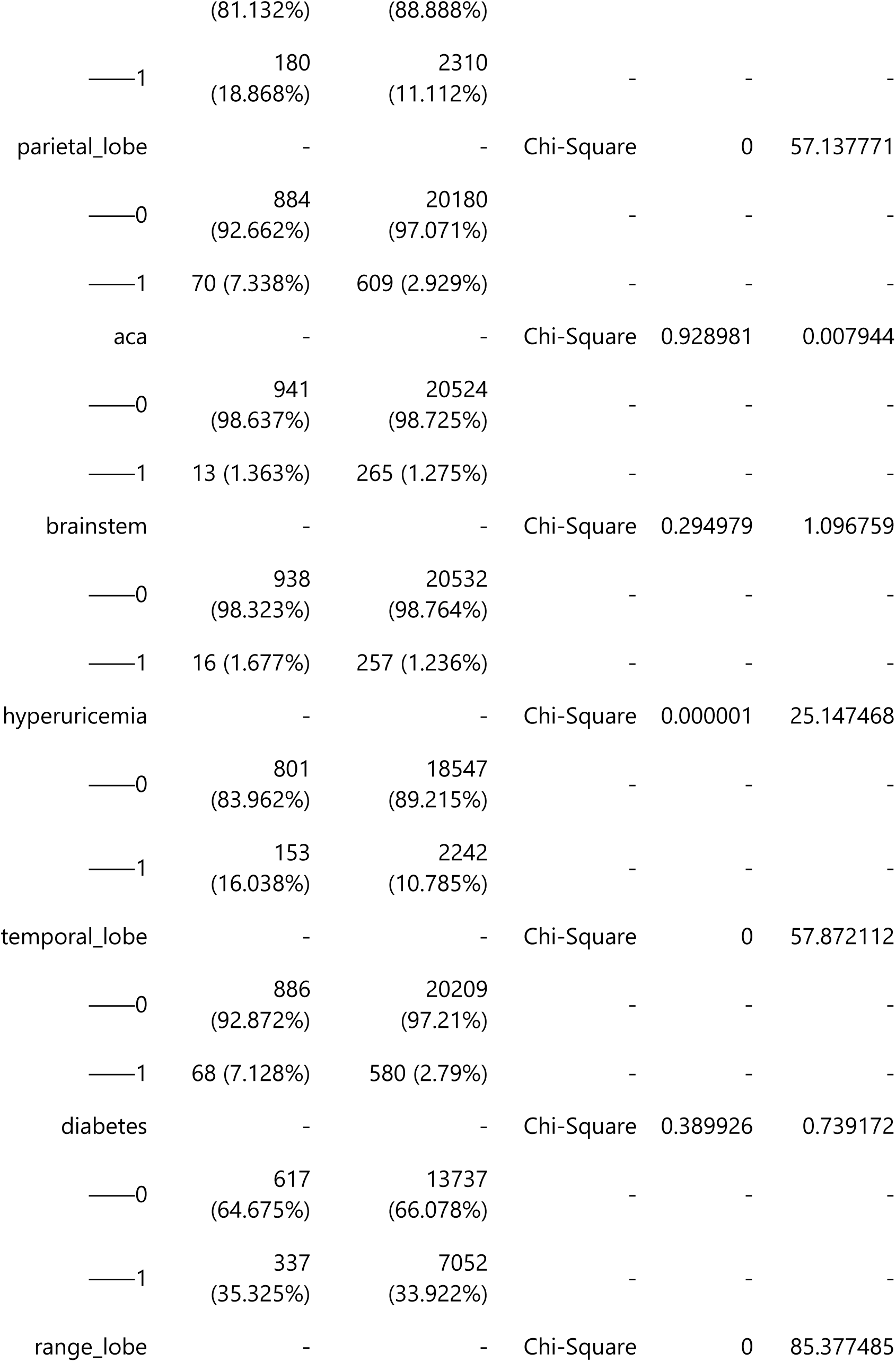
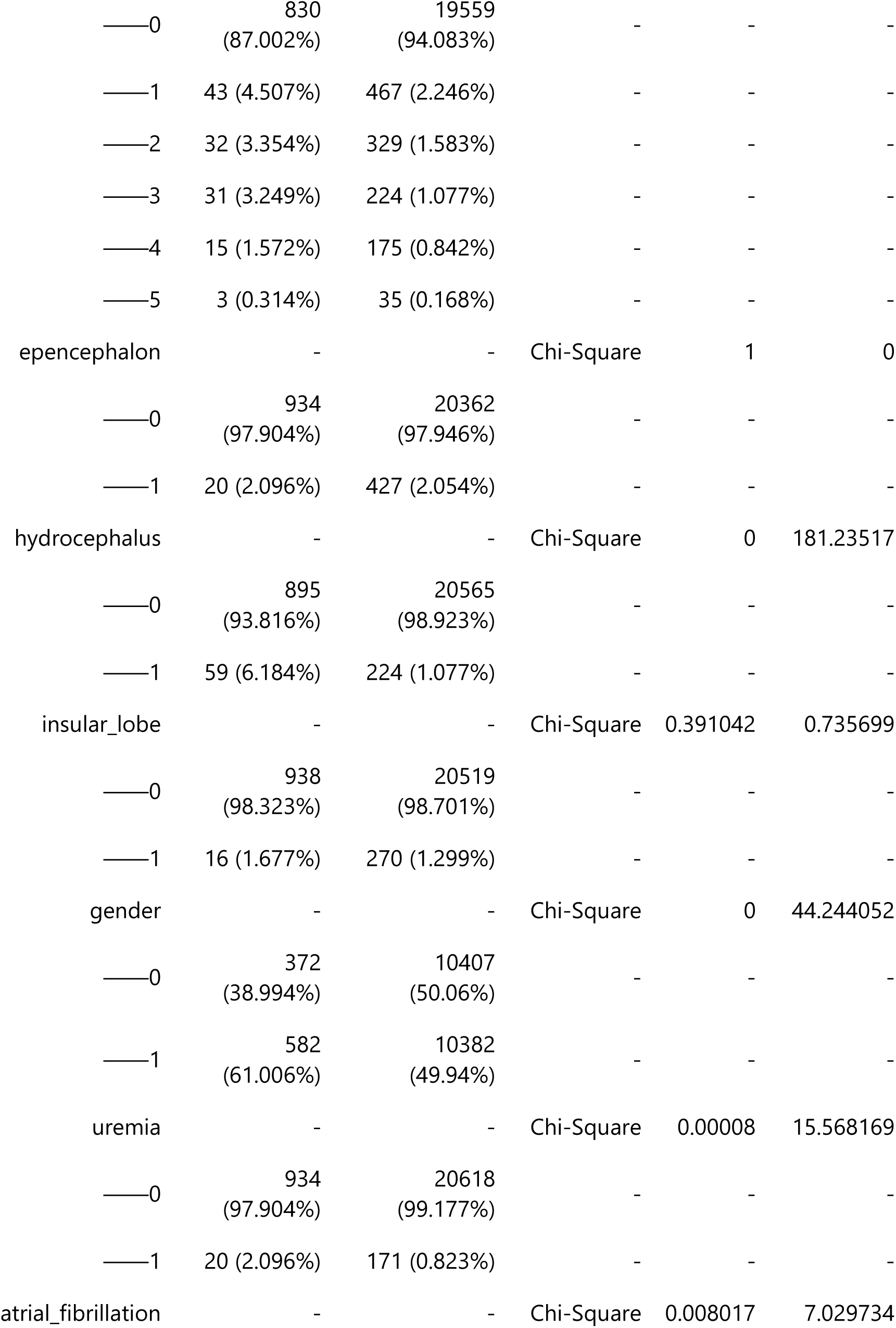
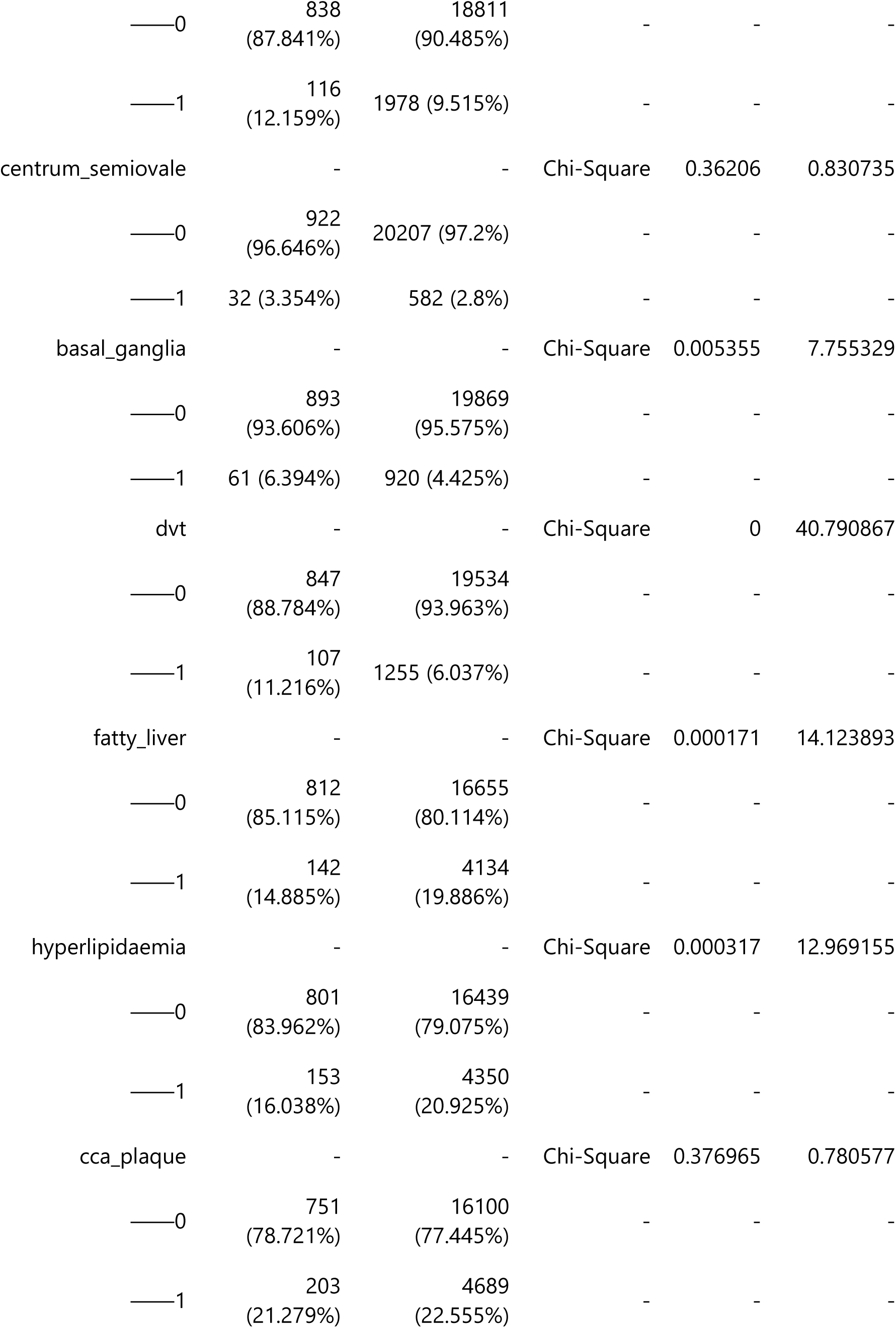
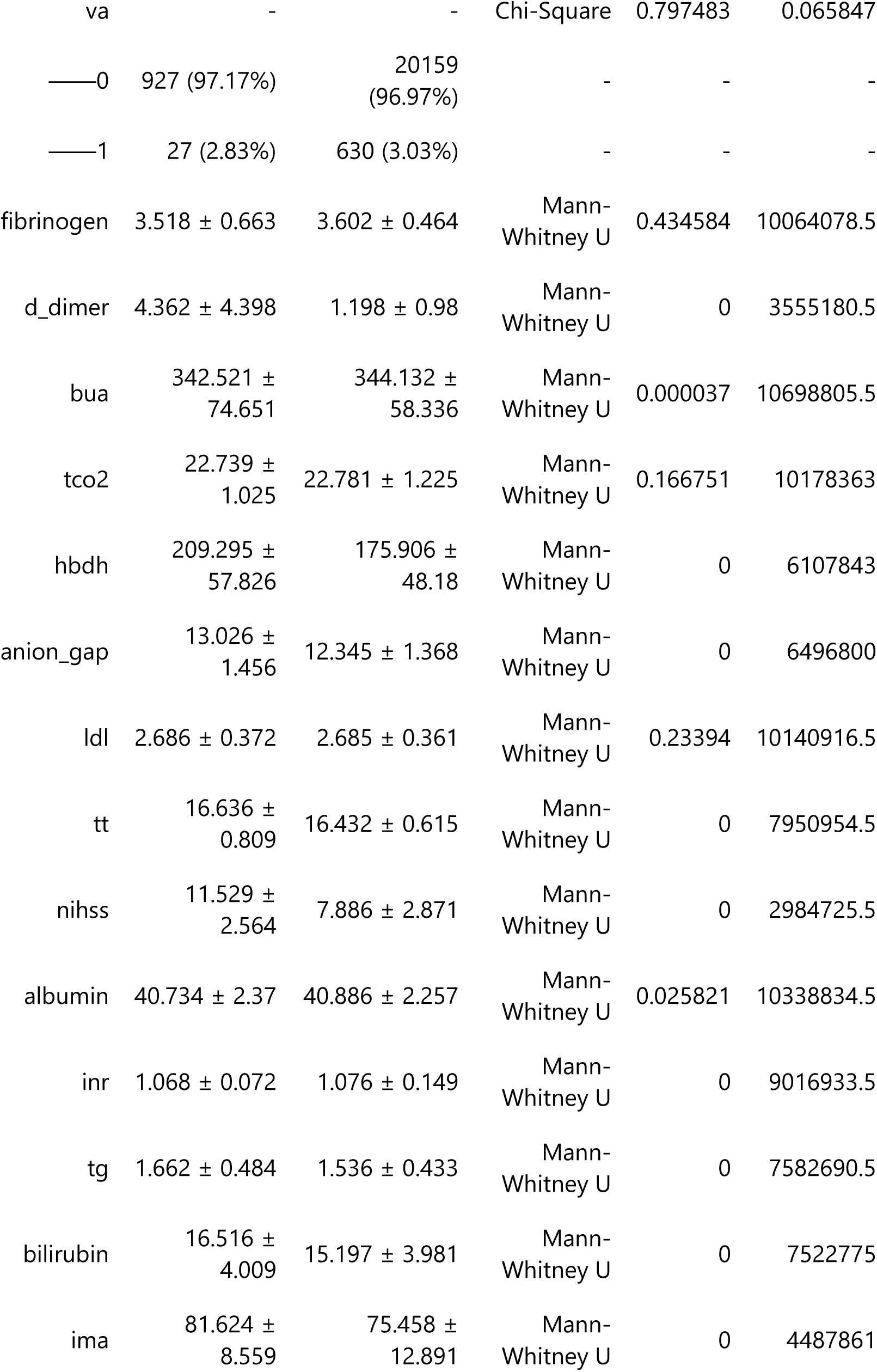
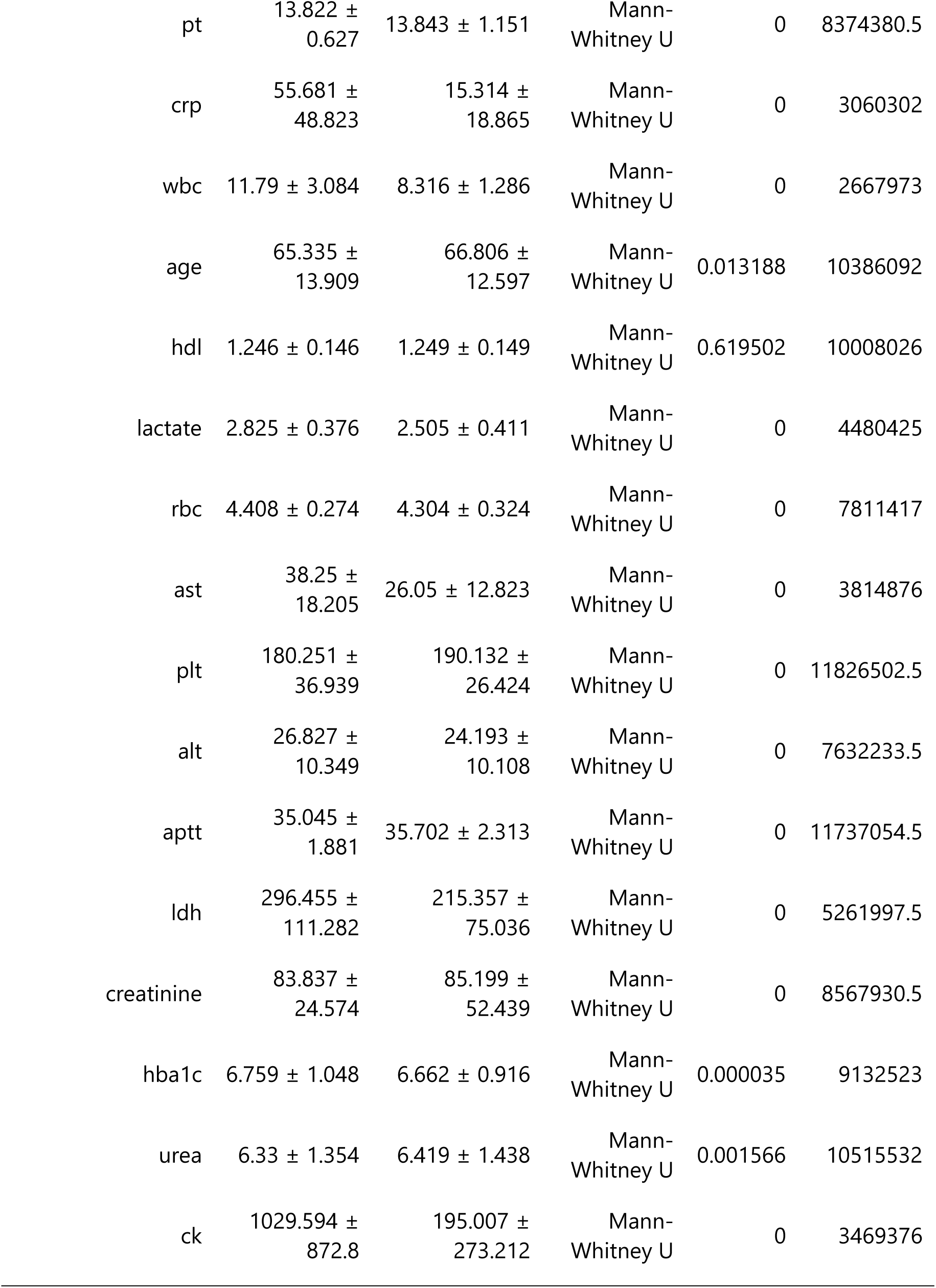
Single factor significant analysis results. This table presents the results of Chi-Square and Mann-Whitney U tests used to evaluate the association of various features with positive and negative samples. Sample Sizes: Positive samples (N=954) and negative samples (N=20,789). Statistical Methods: The Chi-Square test assesses the relationship between categorical variables, while the Mann-Whitney U test compares differences between independent groups for continuous data. P-values: Indicate the significance of the associations, with lower values suggesting stronger evidence against the null hypothesis. Statistical Values: Include counts and percentages of samples for each feature in both groups, along with the calculated statistics for each test.

Statistical analysis indicated that patients with a higher likelihood of developing PSE had complications such as uremia, a history of DVT, atrial fibrillation, hyperuricemia, cerebral hernia, and hydrocephalus. The affected brain regions included the frontal, parietal, occipital, and temporal lobes, as well as the cortex, subcortex, basal ganglia, and hypothalamus. General characteristics included age, gender, and NIHSS score. Laboratory indicators associated with a higher risk of PSE included WBC count, HbA1C, CRP, triglycerides, AST, ALT, bilirubin, urea, uric acid, APTT, PT, D-dimer, CK, CK-MB, LDH, HBDH, IMA, lactate, and anion gap. Additionally, significant p-values were found for fatty liver, coronary heart disease, hyperlipidemia, and HDL, with low or negative values of these indicators linked to a higher risk of secondary complications. The results of the statistical analyses, as well as the univariate and multivariate regression analyses, are detailed in Tables 1, 2, and 3.

**Table 2.**
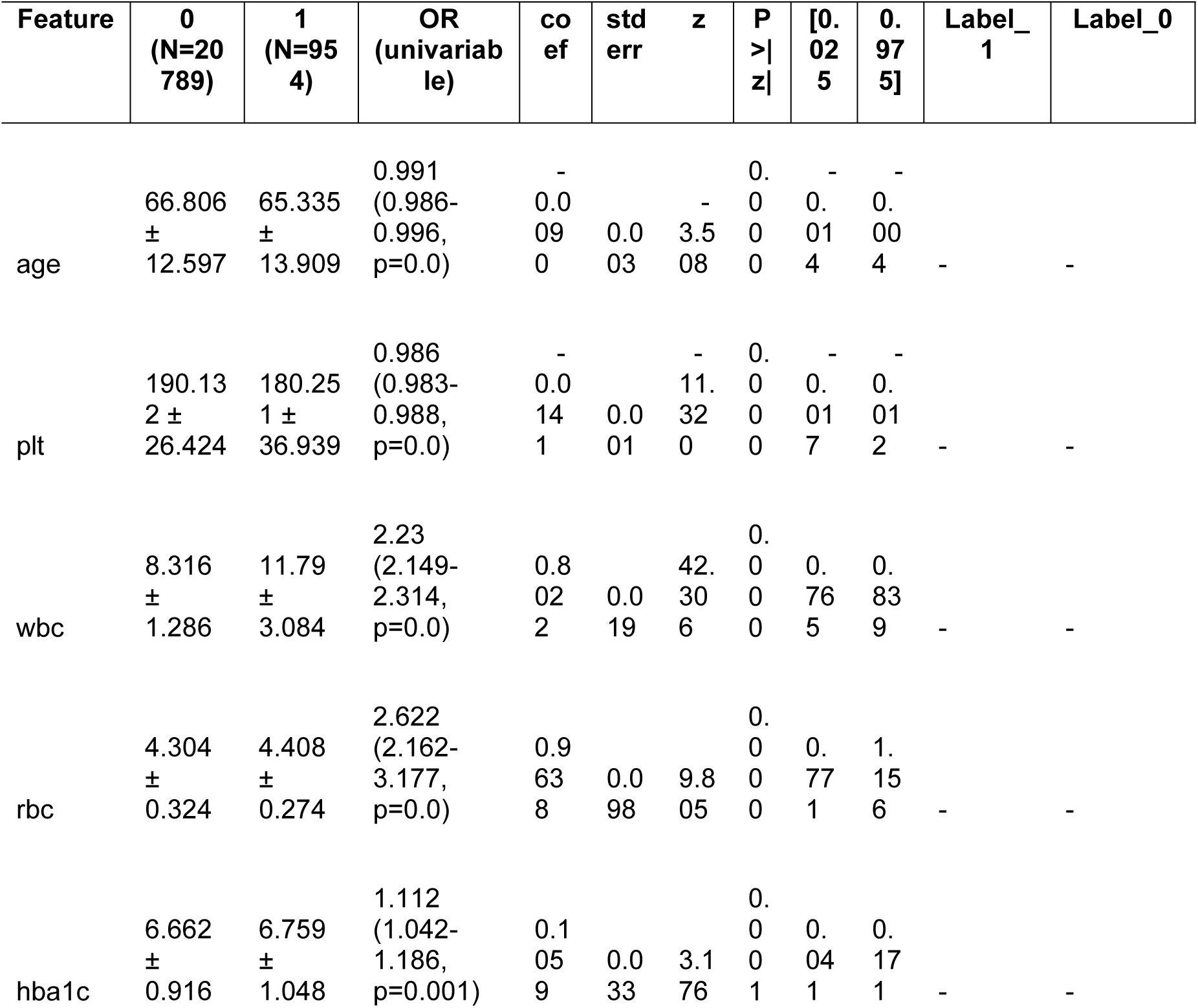
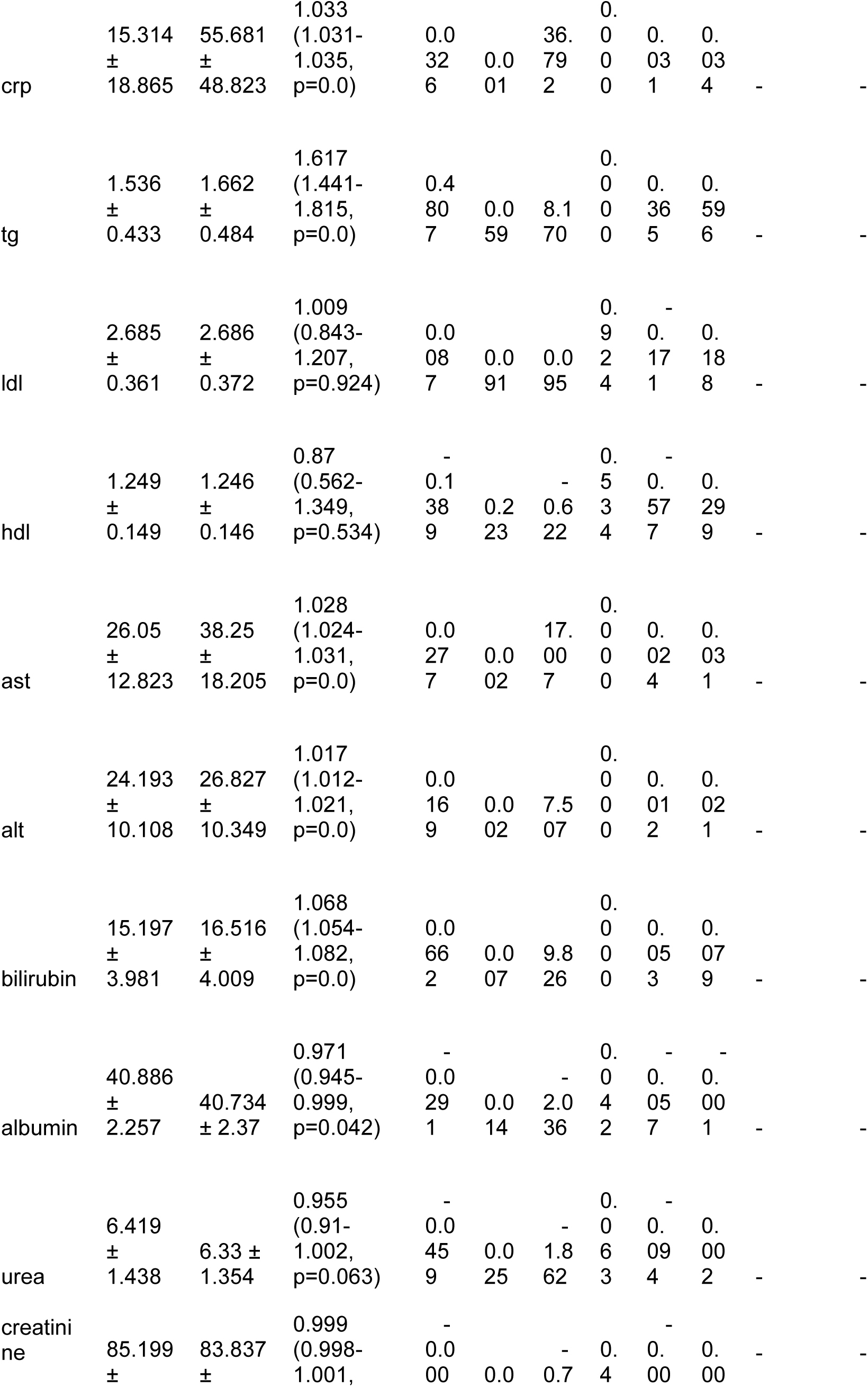
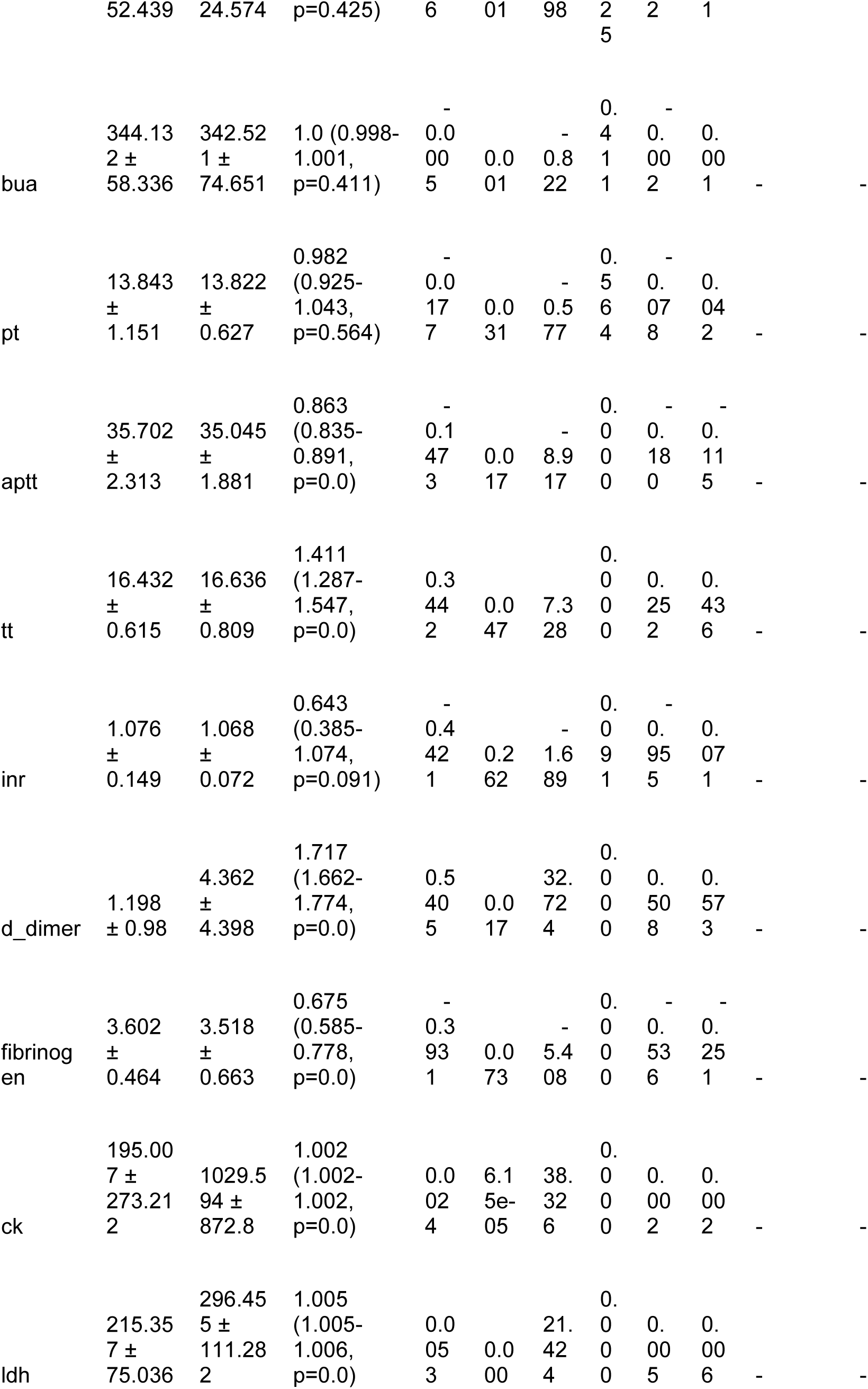
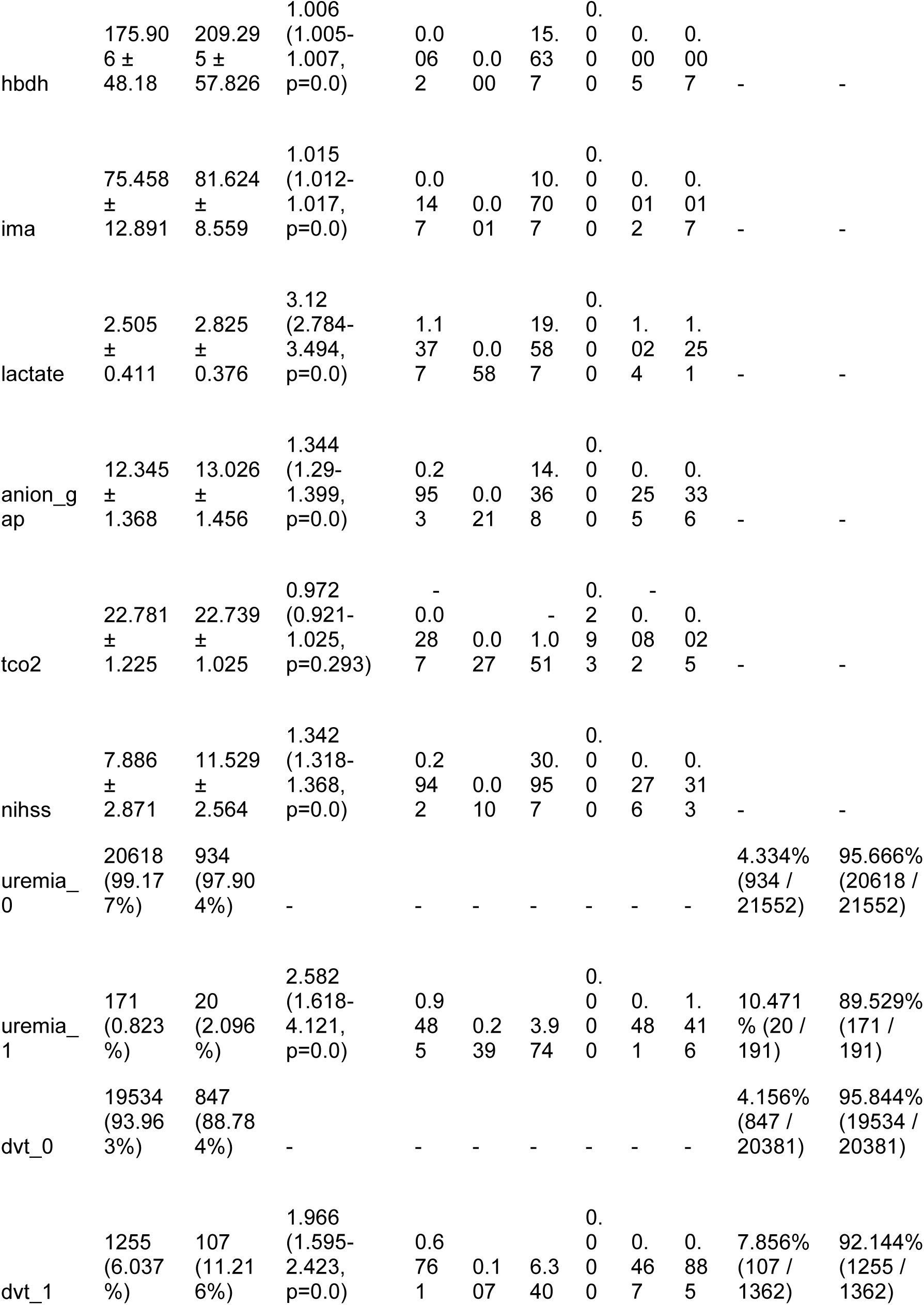
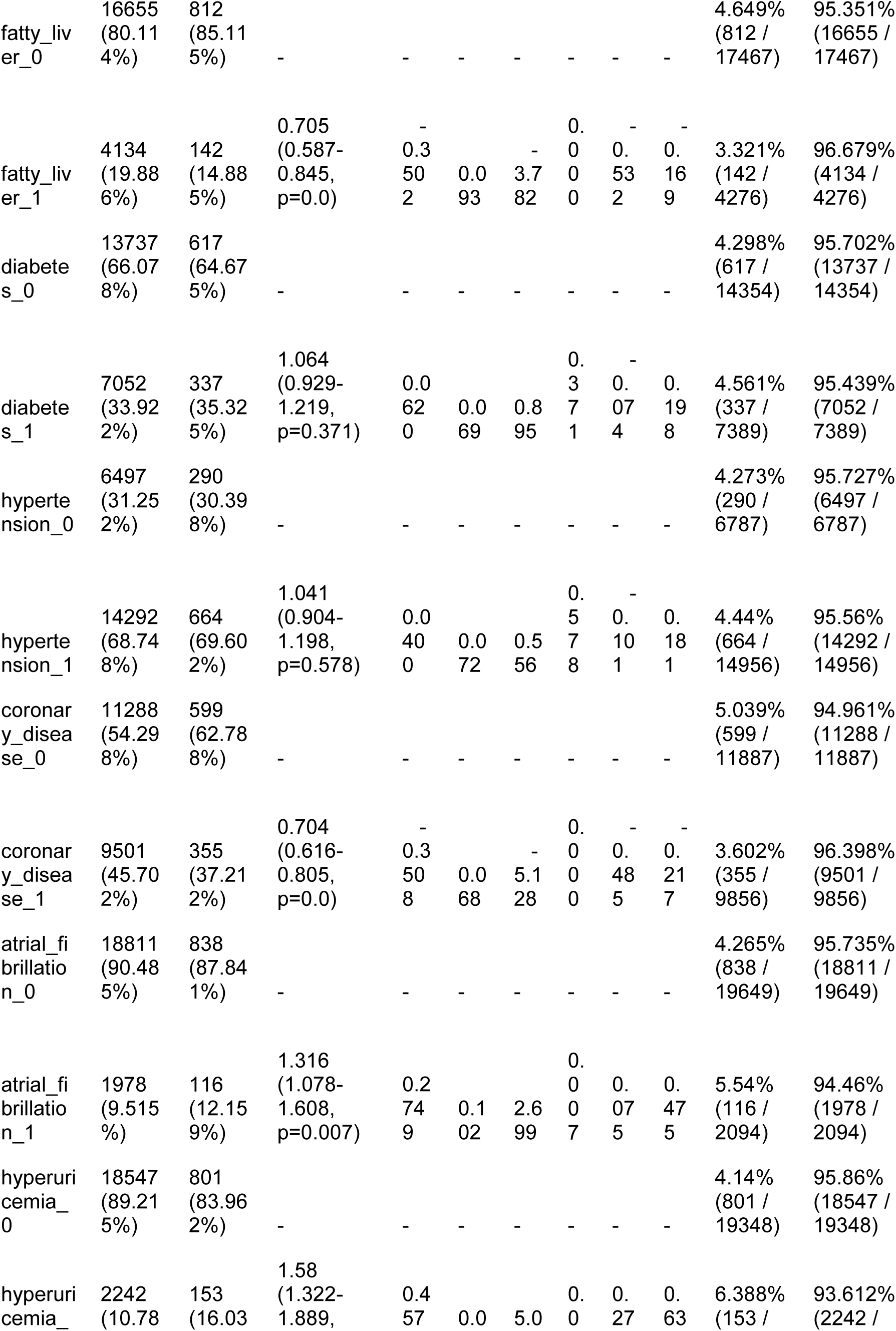
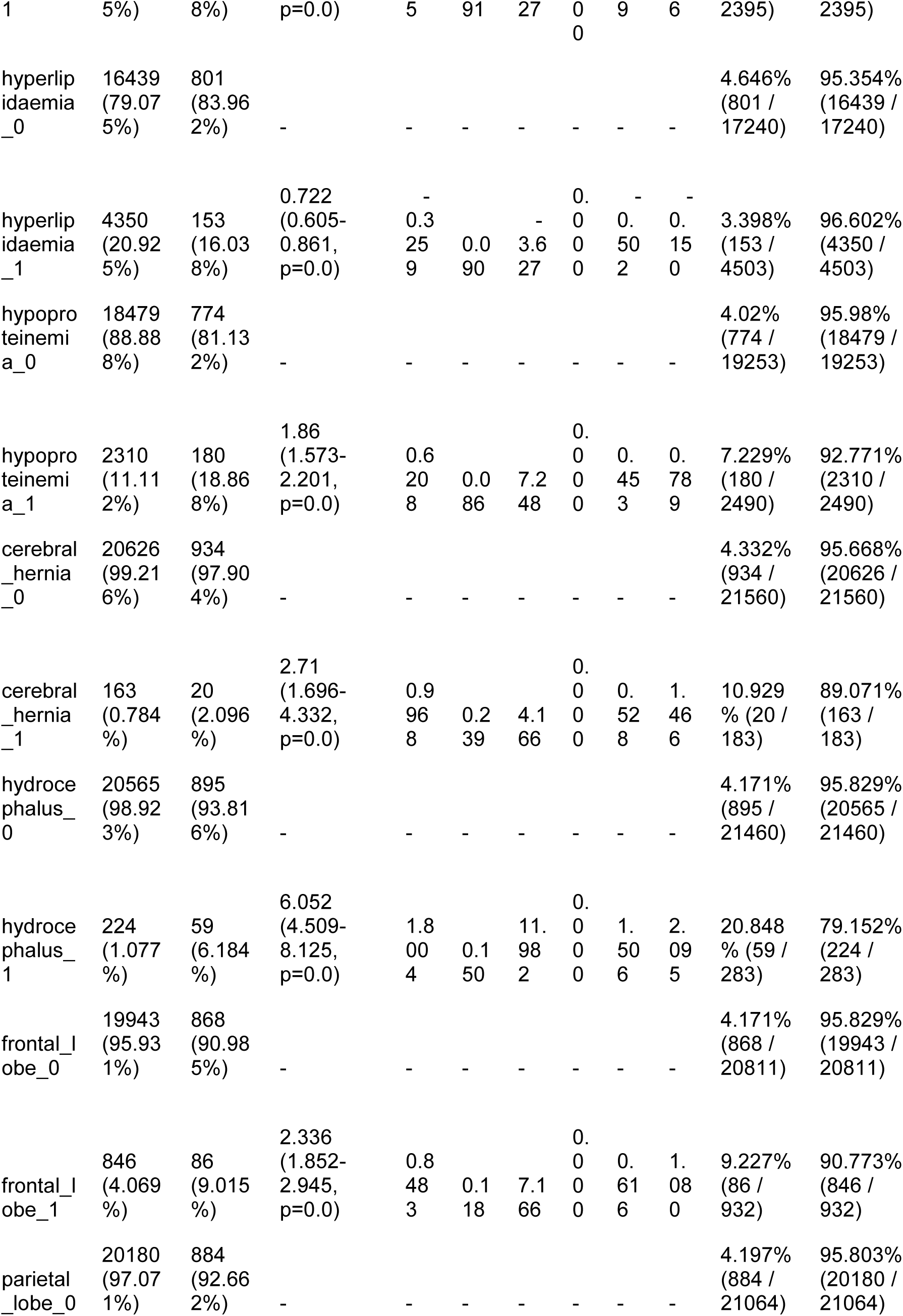
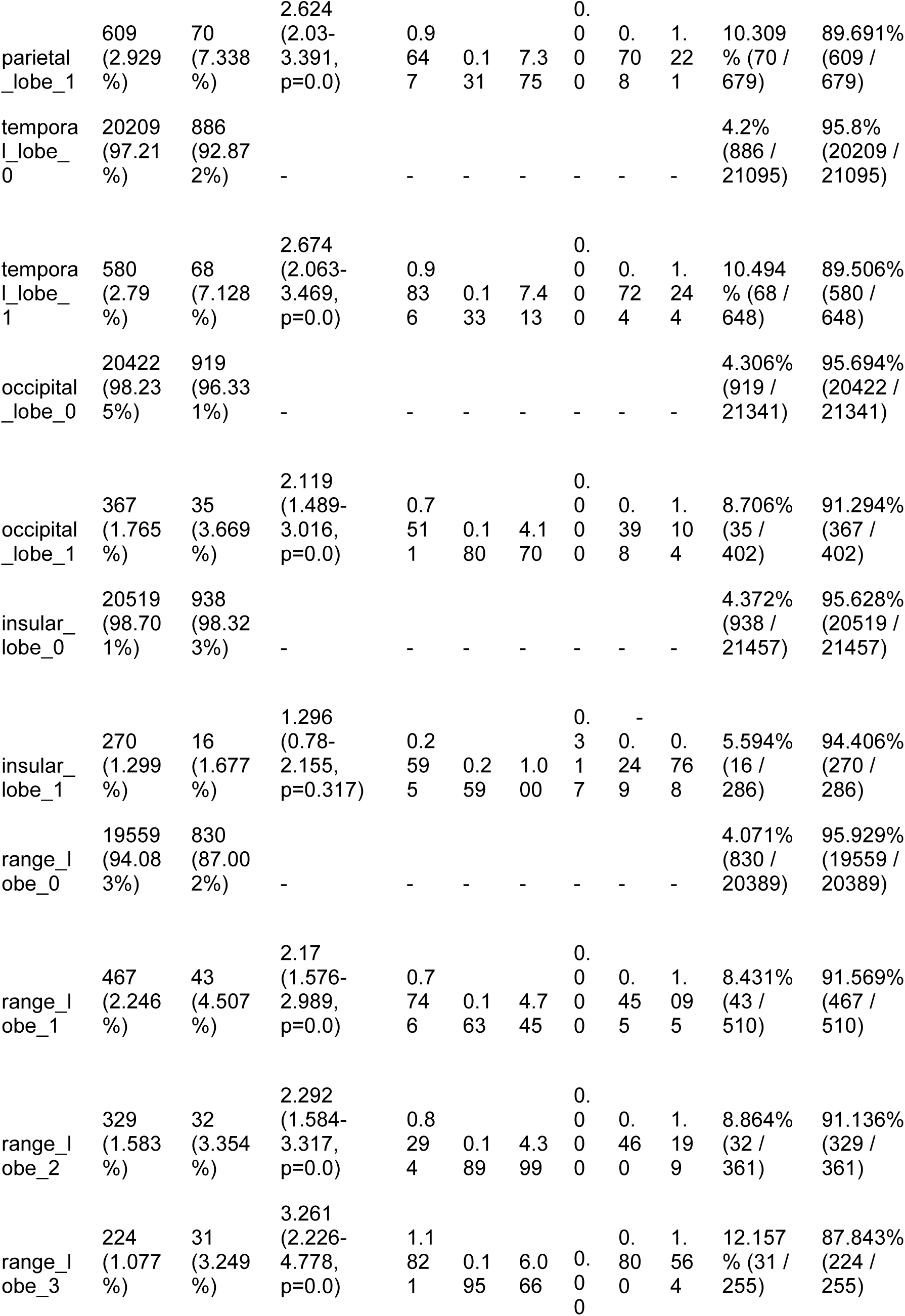
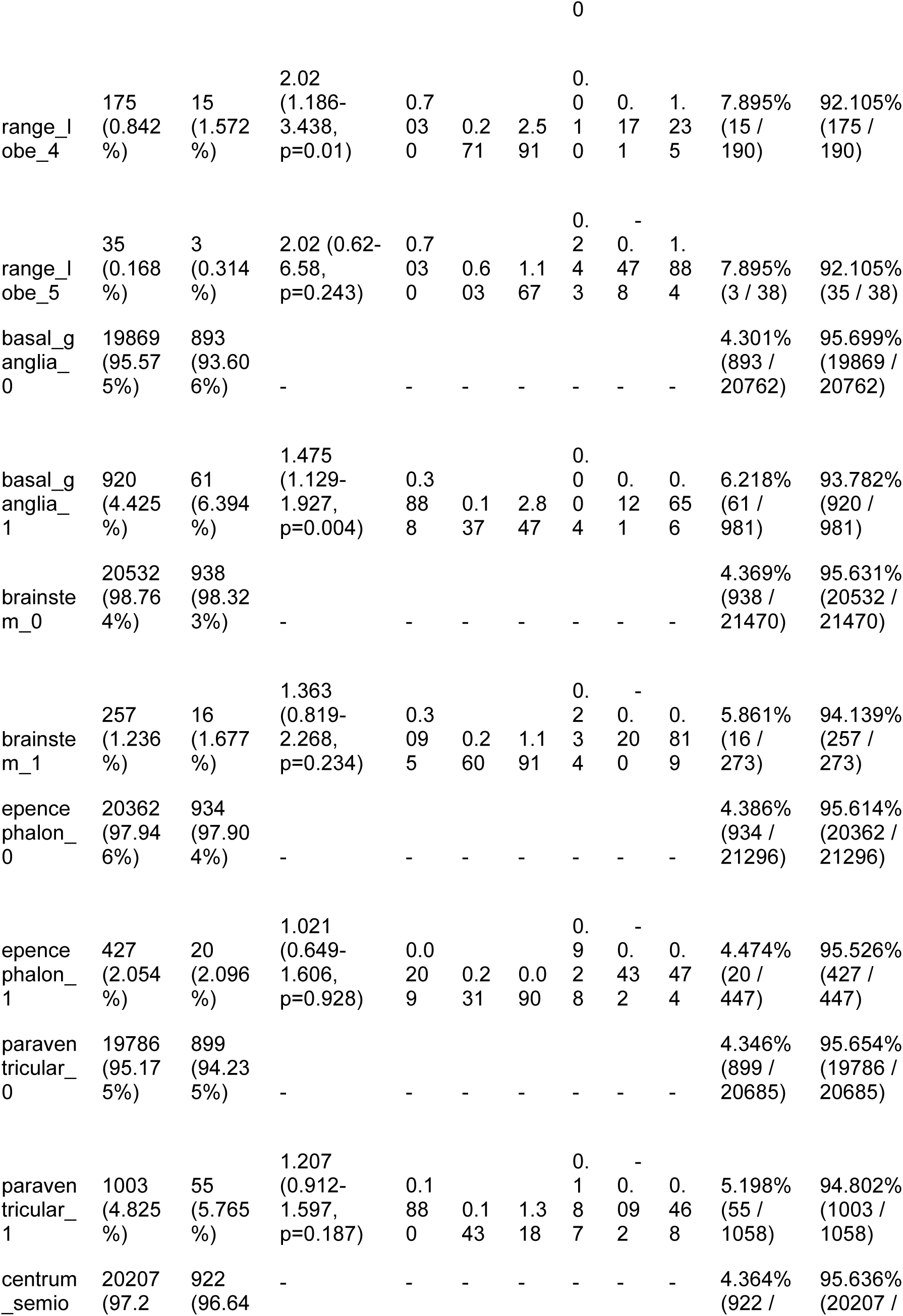
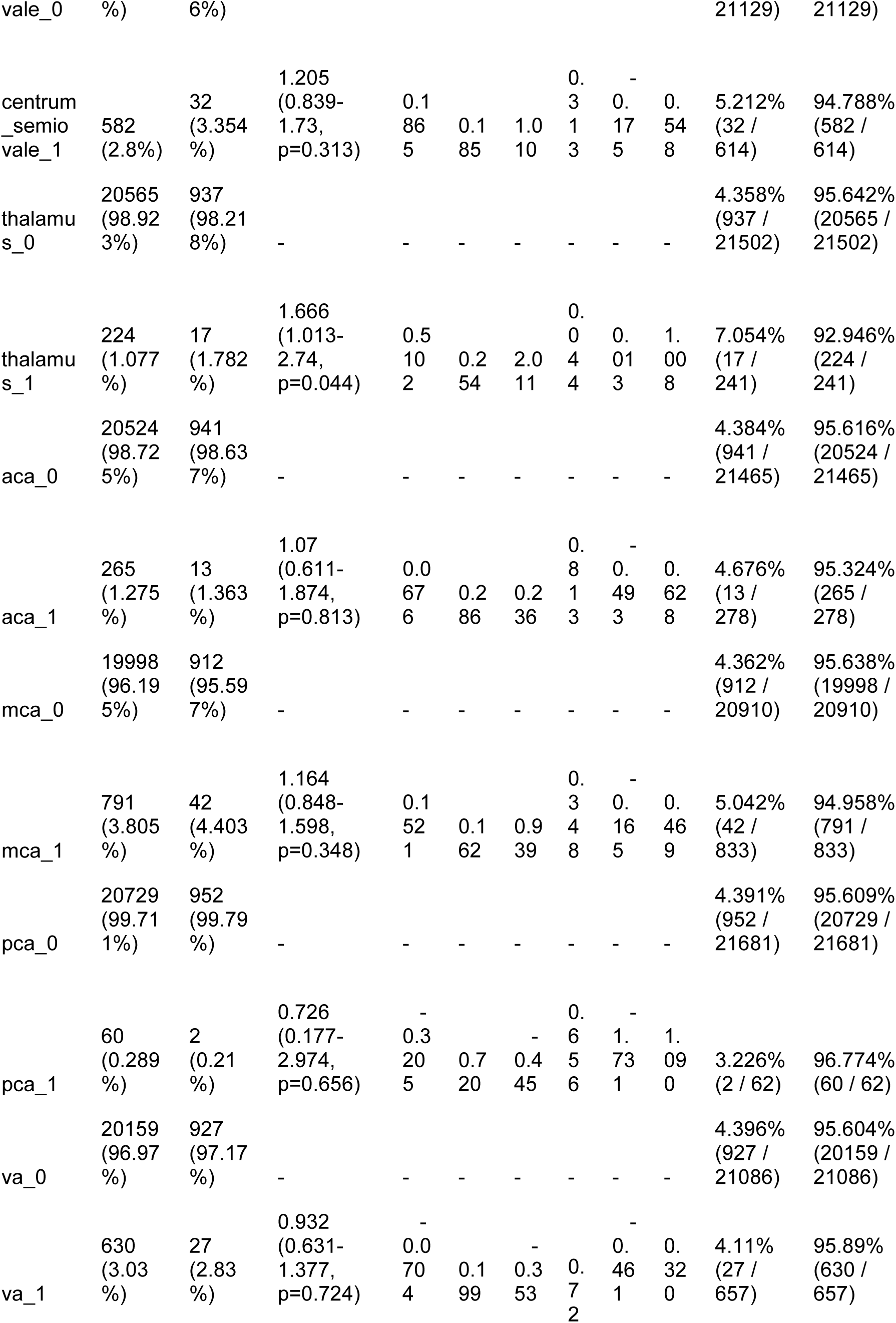
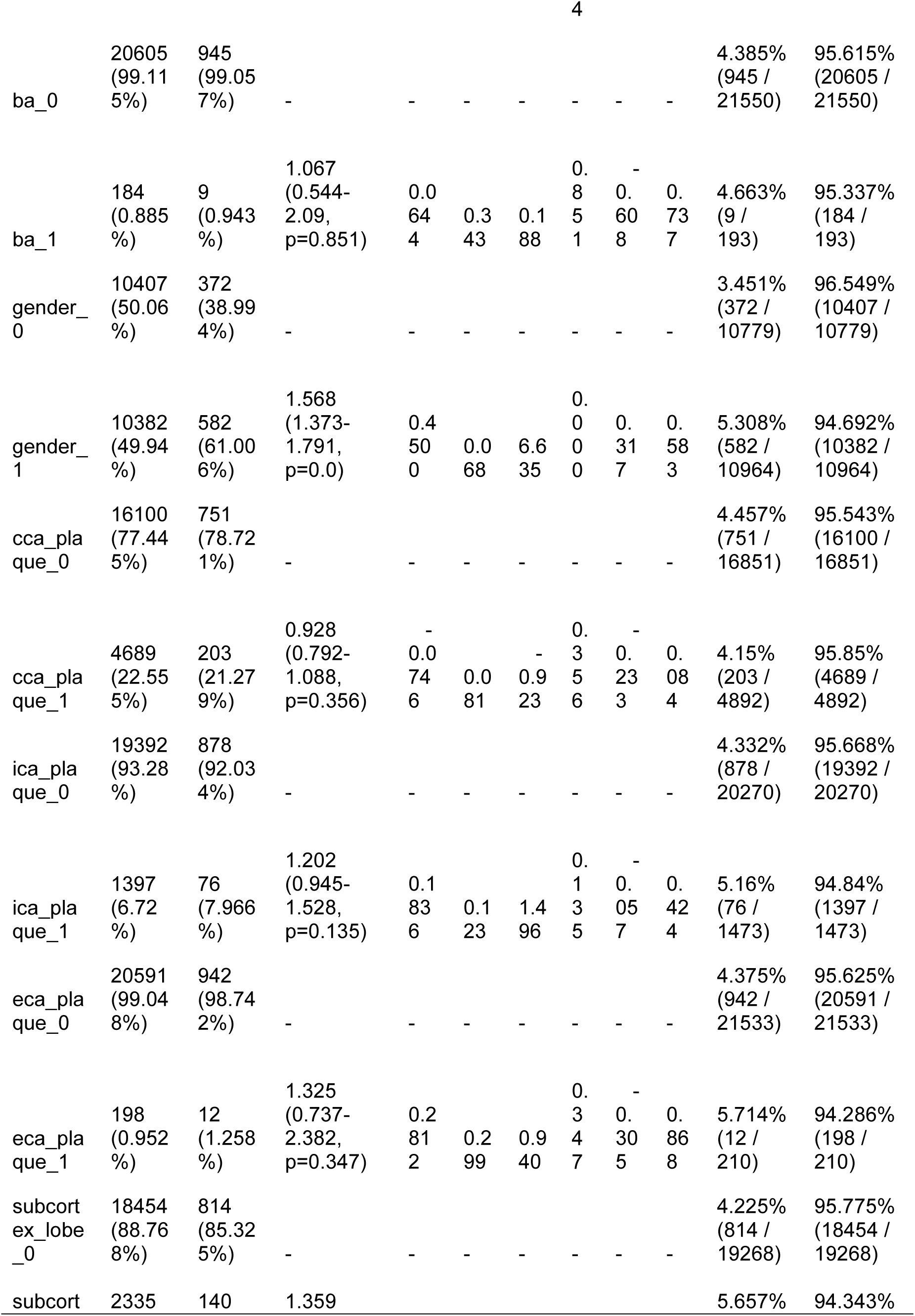
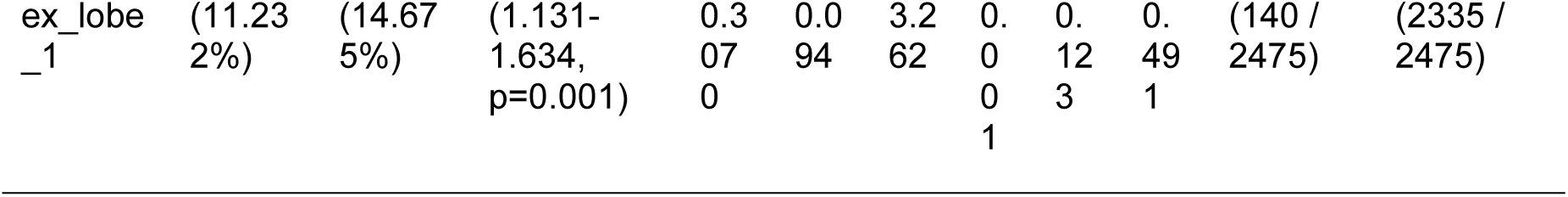
Single Factor Significant Analysis Results. This table presents the results of a single factor significance analysis for various features across two groups of samples: negative samples (0) and positive samples (1). Sample Size: Group 0 (Negative): N = 20,789 Group 1 (Positive): N = 954 Feature Analysis: For each feature, the table includes the mean and standard deviation (±) for both groups, odds ratios (OR) from univariable analysis, coefficients (coef), standard errors (std err), z-scores (z), p-values (P>|z|), and 95% confidence intervals ([0.025, 0.975]). Significance Levels: Features with statistically significant differences are indicated by p-values less than 0.05. An odds ratio greater than 1 suggests an increased risk associated with the feature in the positive group, while an odds ratio less than 1 suggests a decreased risk. Labels: The last two columns provide the proportions of the positive and negative samples for selected features.

### Performance of machine learning models

The relevant performance indicators of the machine learning models are presented in Table 4, while the ROC curves, calibration curve, and decision curve analysis (DCA) are shown in Figure 3. Among all models, tree-based models such as Random Forest (RF), XGBoost, and LightGBM had the highest AUC scores, outperforming other models. Notably, Random Forest had the highest positive predictive value (PPV) at 0.864, which was the most significant metric in our models. Complex machine learning algorithms performed better than traditional logistic regression. The Brier score of the calibration curve was 0.006, and the DCA demonstrated good clinical decision-making benefits, indicating strong practical value. In the external validation cohort, we used RF for predictions, achieving a sensitivity of 0.91 and a PPV of 0.95, confirming the model’s strong predictive capability.

**Table 3.** Multivariable Analysis Results. This table summarizes the results of a multivariable analysis for various features across two groups of samples: negative samples (0) and positive samples (1). Sample Size: Group 0 (Negative): N = 20,789 Group 1 (Positive): N = 954 Feature Analysis: For each feature, the table includes the mean and standard deviation (±) for both groups, odds ratios (OR) from multivariable analysis, coefficients (Coef.), standard errors (Std. Err.), z-scores (z), p-values (P>|z|), and 95% confidence intervals ([0.025, 0.975]). Significance Levels: Features with statistically significant differences are indicated by p-values less than 0.05. An odds ratio greater than 1 indicates an increased risk associated with the feature in the positive group, while an odds ratio less than 1 suggests a decreased risk. Labels: The last column presents the proportions of the positive and negative samples for selected features.

| Feature | 0<br>(N=20789) | 1<br>(N=954) | OR<br>(multivariable) | Coef. | Std. Err. | z | P>\|z\| | [0.025 | 0.975] |
| --- | --- | --- | --- | --- | --- | --- | --- | --- | --- |
| tg | 1.536 ± 0.433 | 1.662 ± 0.484 | 2.458<br>(2.069-2.92,<br>p=0.0) | 0.899 | 0.088 | 10.23 | 0 | 0.727 | 1.071 |
| rbc | 4.304 ± 0.324 | 4.408 ± 0.274 | 4.731<br>(3.274-6.837,<br>p=0.0) | 1.554 | 0.188 | 8.275 | 0 | 1.186 | 1.922 |
| age | 66.806 ± 12.597 | 65.335 ± 13.909 | 1.012<br>(1.004-1.021,<br>p=0.003) | 0.012 | 0.004 | 2.971 | 0.003 | 0.004 | 0.021 |
| ast | 26.05 ± 12.823 | 38.25 ± 18.205 | 1.048 (1.04-1.055,<br>p=0.0) | 0.046 | 0.004 | 12.413 | 0 | 0.039 | 0.054 |
| plt | 190.132 ± 26.424 | 180.251 ± 36.939 | 0.977<br>(0.973-0.98,<br>p=0.0) | -0.024 | 0.002 | -13.375 | 0 | -0.027 | -0.02 |
| alt | 24.193<br>±<br>10.108 | 26.827<br>±<br>10.349 | 0.953<br>(0.942-0.964,<br>p=0.0) | -<br>0.0<br>48 | 0.006 | -<br>8.17<br>7 | 0 | -<br>0.05<br>9 | -<br>0.03<br>6 |
| ima | 75.458<br>±<br>12.891 | 81.624<br>± 8.559 | 1.006<br>(1.001-1.012,<br>p=0.014) | 0.0<br>06 | 0.003 | 2.45<br>3 | 0.0<br>14 | 0.00<br>1 | 0.01<br>2 |
| ldh | 215.35<br>7 ±<br>75.036 | 296.45<br>5 ±<br>111.28<br>2 | 0.984<br>(0.982-0.987,<br>p=0.0) | -<br>0.0<br>16 | 0.001 | -<br>12.9<br>92 | 0 | -<br>0.01<br>8 | -<br>0.01<br>4 |
| tt | 16.432<br>± 0.615 | 16.636<br>± 0.809 | 1.13 (1.009-1.265,<br>p=0.034) | 0.1<br>22 | 0.058 | 2.11<br>6 | 0.0<br>34 | 0.00<br>9 | 0.23<br>5 |
| crp | 15.314<br>±<br>18.865 | 55.681<br>±<br>48.823 | 1.032<br>(1.028-1.036,<br>p=0.0) | 0.0<br>31 | 0.002 | 15.5<br>85 | 0 | 0.02<br>7 | 0.03<br>5 |
| wbc | 8.316 ±<br>1.286 | 11.79 ±<br>3.084 | 2.091<br>(1.985-2.204,<br>p=0.0) | 0.7<br>38 | 0.027 | 27.5<br>83 | 0 | 0.68<br>5 | 0.79 |
| ck | 195.00<br>7 ±<br>273.21<br>2 | 1029.5<br>94 ±<br>872.8 | 1.001<br>(1.001-1.001,<br>p=0.0) | 0.0<br>01 | 0 | 7.86 | 0 | 0.00<br>1 | 0.00<br>1 |
| subcortex_lobe_0 | 18454<br>(88.768<br>%) | 814<br>(85.325<br>%) | - | - | - | - | - | - | - |
| subcortex_lobe_1 | 2335<br>(11.232<br>%) | 140<br>(14.675<br>%) | 1.188<br>(0.827-1.707 ,p=0.3<br>52) | 0.1<br>72 | 0.185 | 0.93 | 0.3<br>52 | -<br>0.19<br>1 | 0.53<br>5 |
| frontal_lobe_0 | 19943<br>(95.931<br>%) | 868<br>(90.985<br>%) | - | - | - | - | - | - | - |
| frontal_lobe_1 | 846<br>(4.069<br>%) | 86<br>(9.015<br>%) | 4.577<br>(1.381-15.17 ,p=0.0<br>13) | 1.5<br>21 | 0.611 | 2.48<br>8 | 0.0<br>13 | 0.32<br>3 | 2.71<br>9 |
| cerebral_hernia_0 | 20626<br>(99.216<br>%) | 934<br>(97.904<br>%) | - | - | - | - | - | - | - |
| cerebral_hernia_1 | 163<br>(0.784<br>%) | 20<br>(2.096<br>%) | 0.846<br>(0.387-1.85 ,p=0.67<br>6) | -<br>0.1<br>67 | 0.399 | -<br>0.41<br>8 | 0.6<br>76 | -<br>0.94<br>9 | 0.61<br>5 |
| thalamus_0 | 20565<br>(98.923<br>%) | 937<br>(98.218<br>%) | - | - | - | - | - | - | - |
| thalamus_1 | 224<br>(1.077<br>%) | 17<br>(1.782<br>%) | 0.669<br>(0.327-1.373 ,p=0.2<br>73) | -<br>0.4<br>01 | 0.366 | -<br>1.09<br>5 | 0.2<br>73 | -<br>1.11<br>9 | 0.31<br>7 |
| occipital_lobe_0 | 20422<br>(98.235<br>%) | 919<br>(96.331<br>%) | - | - | - | - | - | - | - |
| occipital_lobe_1 | 367<br>(1.765<br>%) | 35<br>(3.669<br>%) | 2.172<br>(0.741-6.368 ,p=0.1<br>57) | 0.7<br>76 | 0.549 | 1.41<br>4 | 0.1<br>57 | -0.3 | 1.85<br>1 |
| coronary_disease_0 | 11288<br>(54.298<br>%) | 599<br>(62.788<br>%) | - | - | - | - | - | - | - |
| coronary_disease_1 | 9501<br>(45.702<br>%) | 355<br>(37.212<br>%) | 1.408<br>(1.151-1.724 ,p=0.0<br>01) | 0.3<br>42 | 0.103 | 3.32<br>2 | 0.0<br>01 | 0.14 | 0.54<br>5 |
| hypoproteinaemia_0 | 18479<br>(88.888<br>%) | 774<br>(81.132<br>%) | - | - | - | - | - | - | - |
| hypoproteinaemia_1 | 2310<br>(11.112<br>%) | 180<br>(18.868<br>%) | 1.183 (0.9-1.554 ,p=0.2<br>28) | 0.1<br>68 | 0.139 | 1.20<br>6 | 0.2<br>28 | -<br>0.10<br>5 | 0.44<br>1 |
| parietal_lobe_0 | 20180<br>(97.071<br>%) | 884<br>(92.662<br>%) | - | - | - | - | - | - | - |
| parietal_lobe_1 | 609<br>(2.929<br>%) | 70<br>(7.338<br>%) | 6.939<br>(2.253-21.375 ,p=0.<br>001) | 1.9<br>37 | 0.574 | 3.37<br>5 | 0.0<br>01 | 0.81<br>2 | 3.06<br>2 |
| hyperuricemia_0 | 18547<br>(89.215<br>%) | 801<br>(83.962<br>%) | - | - | - | - | - | - | - |
| hyperuricemia<br>_1 | 2242<br>(10.785<br>%) | 153<br>(16.038<br>%) | 0.938<br>(0.691-1.275 ,p=0.6<br>84) | -<br>0.0<br>64 | 0.156 | -<br>0.40<br>7 | 0.6<br>84 | -0.37 | 0.24<br>3 |
| temporal_lobe<br>_0 | 20209<br>(97.21<br>%) | 886<br>(92.872<br>%) | - | - | - | - | - | - | - |
| temporal_lobe<br>_1 | 580<br>(2.79%) | 68<br>(7.128<br>%) | 5.242<br>(1.548-17.752 ,p=0.<br>008) | 1.6<br>57 | 0.622 | 2.66<br>2 | 0.0<br>08 | 0.43<br>7 | 2.87<br>6 |
| range_lobe_0 | 19559<br>(94.083<br>%) | 830<br>(87.002<br>%) | - | - | - | - | - | - | - |
| range_lobe_1 | 467<br>(2.246<br>%) | 43<br>(4.507<br>%) | 0.359<br>(0.111-1.159 ,p=0.0<br>87) | -<br>1.0<br>25 | 0.598 | -<br>1.71<br>3 | 0.0<br>87 | -<br>2.19<br>7 | 0.14<br>7 |
| range_lobe_2 | 329<br>(1.583<br>%) | 32<br>(3.354<br>%) | 0.084 (0.01-0.703 ,p=0.0<br>22) | -<br>2.4<br>79 | 1.085 | -<br>2.28<br>5 | 0.0<br>22 | -<br>4.60<br>5 | -<br>0.35<br>2 |
| range_lobe_3 | 224<br>(1.077<br>%) | 31<br>(3.249<br>%) | 0.011 (0.0-0.231 ,p=0.0<br>04) | -<br>4.5<br>42 | 1.569 | -<br>2.89<br>5 | 0.0<br>04 | -<br>7.61<br>7 | -<br>1.46<br>7 |
| range_lobe_4 | 175<br>(0.842<br>%) | 15<br>(1.572<br>%) | 0.001 (0.0-0.057 ,p=0.0<br>01) | -<br>6.6<br>66 | 1.943 | -3.43 | 0.0<br>01 | -<br>10.4<br>75 | -<br>2.85<br>7 |
| range_lobe_5 | 35<br>(0.168<br>%) | 3<br>(0.314<br>%) | 0.001 (0.0-0.115 ,p=0.0<br>04) | -<br>6.5<br>86 | 2.259 | -<br>2.91<br>5 | 0.0<br>04 | -<br>11.0<br>14 | -<br>2.15<br>9 |
| hydrocephalus<br>_0 | 20565<br>(98.923<br>%) | 895<br>(93.816<br>%) | - | - | - | - | - | - | - |
| hydrocephalus<br>_1 | 224<br>(1.077<br>%) | 59<br>(6.184<br>%) | 3.251<br>(1.939-5.451 ,p=0.0<br>) | 1.1<br>79 | 0.264 | 4.47<br>1 | 0 | 0.66<br>2 | 1.69<br>6 |
| gender_0 | 10407<br>(50.06 | 372<br>(38.994 | - | - | - | - | - | - | - |
|  | %) | %) |  |  |  |  |  |  |  |
| gender_1 | 10382<br>(49.94<br>%) | 582<br>(61.006<br>%) | 0.572<br>(0.454-0.72 ,p=0.0) | -<br>0.5<br>58 | 0.117 | -<br>4.75<br>3 | 0 | -<br>0.78<br>9 | -<br>0.32<br>8 |
| uremia_0 | 20618<br>(99.177<br>%) | 934<br>(97.904<br>%) | - | - | - | - | - | - | - |
| uremia_1 | 171<br>(0.823<br>%) | 20<br>(2.096<br>%) | 1.979<br>(1.098-3.564 ,p=0.0<br>23) | 0.6<br>82 | 0.3 | 2.27<br>3 | 0.0<br>23 | 0.09<br>4 | 1.27<br>1 |
| atrial_fibrillatio<br>n_0 | 18811<br>(90.485<br>%) | 838<br>(87.841<br>%) | - | - | - | - | - | - | - |
| atrial_fibrillatio<br>n_1 | 1978<br>(9.515<br>%) | 116<br>(12.159<br>%) | 1.446<br>(1.087-1.923 ,p=0.0<br>11) | 0.3<br>69 | 0.145 | 2.53<br>4 | 0.0<br>11 | 0.08<br>4 | 0.65<br>4 |
| basal_ganglia_<br>0 | 19869<br>(95.575<br>%) | 893<br>(93.606<br>%) | - | - | - | - | - | - | - |
| basal_ganglia_<br>1 | 920<br>(4.425<br>%) | 61<br>(6.394<br>%) | 1.024<br>(0.642-1.633 ,p=0.9<br>21) | 0.0<br>24 | 0.238 | 0.09<br>9 | 0.9<br>21 | -<br>0.44<br>3 | 0.49 |
| dvt_0 | 19534<br>(93.963<br>%) | 847<br>(88.784<br>%) | - | - | - | - | - | - | - |
| dvt_1 | 1255<br>(6.037<br>%) | 107<br>(11.216<br>%) | 1.254<br>(0.922-1.706 ,p=0.1<br>49) | 0.2<br>27 | 0.157 | 1.44<br>3 | 0.1<br>49 | -<br>0.08<br>1 | 0.53<br>4 |
| hyperlipidaemi<br>a_0 | 16439<br>(79.075<br>%) | 801<br>(83.962<br>%) | - | - | - | - | - | - | - |
| hyperlipidaemi<br>a_1 | 4350<br>(20.925<br>%) | 153<br>(16.038<br>%) | 0.825<br>(0.646-1.052 ,p=0.1<br>21) | -<br>0.1<br>93 | 0.124 | -<br>1.55<br>2 | 0.1<br>21 | -<br>0.43<br>7 | 0.05<br>1 |
| fatty_liver_0 | 16655<br>(80.114 | 812<br>(85.115 | - | - | - | - | - | - | - |
|  | %) | %) |  |  |  |  |  |  |  |
| fatty_liver_1 | 4134<br>(19.886<br>%) | 142<br>(14.885<br>%) | 0.759 (0.59-0.978 ,p=0.0<br>33) | -<br>0.2<br>75 | 0.129 | -<br>2.13<br>5 | 0.0<br>33 | -<br>0.52<br>8 | -<br>0.02<br>3 |

**Table 4.** Model Performance Evaluation Results. This table presents the performance evaluation metrics for various machine learning models, including AUC, Accuracy, Sensitivity (Recall), Specificity, F1-score, Positive Predictive Value (PPV/Precision), and Negative Predictive Value (NPV) AUC: Area Under the Curve, indicating the model’s ability to distinguish between positive and negative samples; values closer to 1 indicate better performance. Accuracy: The proportion of correctly classified samples among the total samples. Sensitivity/Recall: The proportion of correctly identified positive samples out of all actual positive samples. Specificity: The proportion of correctly identified negative samples out of all actual negative samples. F1-score: The harmonic mean of precision and recall, considering both the accuracy and completeness of the model. Positive Predictive Value (PPV/Precision): The proportion of correctly identified positive samples among all samples predicted as positive. Negative Predictive Value (NPV): The proportion of correctly identified negative samples among all samples predicted as negative.

| Model | AUC | Accuracy | Sensitivity/Recall | Specificity | F1-score | PPV/precision |
| --- | --- | --- | --- | --- | --- | --- |
| LR | 0.967 \| 0.973 | 0.928 \| 0.927 | 0.920 \| 0.929 | 0.928 \| 0.927 | 0.530 \| 0.524 | 0.373 \| 0.365 |
| NB | 0.903 \| 0.909 | 0.938 \| 0.936 | 0.634 \| 0.662 | 0.952 \| 0.949 | 0.474 \| 0.472 | 0.378 \| 0.367 |
| DT | 0.997 \| 0.906 | 0.993 \| 0.970 | 1.000 \| 0.836 | 0.993 \| 0.976 | 0.930 \| 0.706 | 0.870 \| 0.610 |
| GB | 0.998 \| 0.992 | 0.987 \| 0.980 | 0.976 \| 0.900 | 0.988 \| 0.983 | 0.871 \| 0.794 | 0.786 \| 0.711 |
| RF | 1.000 \| 0.996 | 0.997 \| 0.989 | 1.000 \| 0.883 | 0.997 \| 0.994 | 0.967 \| 0.873 | 0.936 \| 0.864 |
| MLP | 0.996 \| 0.984 | 0.977 \| 0.972 | 0.975 \| 0.932 | 0.977 \| 0.974 | 0.790 \| 0.744 | 0.664 \| 0.619 |
| XGB | 1.000 \| 0.996 | 0.996 \| 0.988 | 1.000 \| 0.897 | 0.996 \| 0.992 | 0.961 \| 0.867 | 0.926 \| 0.840 |
| LGBM | 1.000 \| 0.996 | 0.997 \| 0.989 | 1.000 \| 0.886 | 0.997 \| 0.993 | 0.970 \| 0.869 | 0.941 \| 0.853 |
| KNN | 0.997 \| 0.955 | 0.965 \| 0.955 | 0.999 \| 0.890 | 0.964 \| 0.958 | 0.717 \| 0.631 | 0.560 \| 0.489 |

### Analysis of SHAP risk factors

Figure 4 shows the SHAP (Shapley Additive Explanations) values, individual decision attempts, and overall decision curves. Among general characteristics, females had a higher rate of PSE. A higher NIHSS score was associated with a higher incidence of PSE. Additionally, elevated values of WBC count, D-dimer, CRP, AST, CK-MB, HbA1c, bilirubin, TCO2, and LDH at admission were linked to a greater likelihood of developing PSE. Conversely, lower levels of HBDH, PLT, and APTT were also associated with a higher probability of PSE. The specific brain regions affected did not have a significant individual effect on the overall outcome. Among complications, hypertension was more strongly associated with PSE development, while other conditions, such as coronary heart disease, diabetes, hyperlipidemia, and fatty liver, were less likely to be related to the outcome. We used the force plot of the first patient to illustrate how different features influenced the prediction. In this case, a prolonged APTT time contributed the most to PSE, followed by elevated AST levels, while a low NIHSS score contributed negatively to the final result. The decision plot aggregated model decisions to show how complex models arrived at their predictions.

## Discussion

Our study used comprehensive clinical, imaging, and laboratory data from stroke patients to develop a predictive model using machine learning algorithms. This model achieved an AUC score above 0.95, demonstrating more accurate predictions compared to traditional statistical methods. Our research revealed that tree-based ensemble models provided superior predictive performance, especially when handling large datasets with high-dimensional features.

During the modeling process, due to the extreme imbalance between negative and positive samples, we applied the SMOTEENN technique to resample the dataset, improving the performance of the machine learning models. Through SHAP analysis, we conducted interpretability assessments of the model and identified the importance of different features.

In our study, age and NIHSS scores were treated as continuous variables. We found that female patients, older individuals, and those with higher NIHSS scores were more likely to develop PSE, consistent with recent studies. Higher NIHSS scores, indicating more severe strokes, significantly increased the risk of complications, second only to white blood cell (WBC) count and D-dimer in our model [5][19][10][20]. However, there are differing views on the effect of age. Some studies [5][21] suggest that age below 65 is a high-risk factor, which aligns with our findings, while other studies [22] have found that advanced age is the key factor. Yamada et al. [21] also agreed with our study, indicating that female patients have a higher risk of complications. On the other hand, Waafi et al. [10] reported that male patients are 3.325 times more likely to develop complications, which contradicts our findings.

Previous research has shown that patients with diabetes, dyslipidemia, hypertension, depression, or dementia are at higher risk of developing vascular epilepsy [12]. In our study, statistical analysis and multiple machine learning (ML) models examined the relationship between comorbidities and complications. We found that patients with coronary heart disease, diabetes, fatty liver, hyperlipidemia, or large artery stenosis or plaques (CCA and ICA) were less likely to develop epilepsy. According to the TOAST classification, ischemic stroke is divided into five categories: large artery atherosclerosis, cardioembolism, small vessel occlusion, other determined etiology, and undetermined etiology. Patients with multiple comorbidities often fall into the large artery atherosclerosis and cardioembolism categories, which are more clearly defined and easier to treat, resulting in a lower likelihood of epilepsy. In contrast, strokes of undetermined etiology tend to have worse prognoses and are more likely to lead to epilepsy. Among patients with diabetes, higher HbA1c levels indicate poor blood sugar control and a higher risk of complications. Patients with better control of their blood sugar have a lower overall risk of developing complications.

Alain et al. found that cortical infarction is more likely to lead to epilepsy in patients hospitalized with anterior circulation ischemic stroke [23]. Lin et al. found that factors such as cortical involvement and intracerebral hemorrhage volume increase the likelihood of PSE, which is consistent with our findings [8]. Al-Sahli et al. also suggested that cortical brain injury and large-area lesions raise the risk of PSE [5][21]. In our study, statistics showed that both cortical and subcortical involvement increased the likelihood of PSE, but these regions had less influence compared to other features and were not selected in the LASSO regression.

Previous studies have identified acute infection as a risk factor for ischemic stroke [24]. C-reactive protein (CRP) reflects inflammation levels and is an independent prognostic factor [25]. In our study, both regression and SHAP analysis indicated that WBC count had a significant impact among routine blood test parameters, even surpassing the NIHSS score in SHAP analysis. A high WBC count may indicate severe inflammation or infection, as well as increased blood viscosity, making patients more prone to secondary complications. In general, a high red blood cell count and low platelet count also contributed to an increased risk of complications.

A large-scale study on Chinese individuals found a negative correlation between plasma high-density lipoprotein cholesterol (HDL-C) levels and the risk of ischemic stroke, a weak positive correlation between plasma triglyceride (TG) levels and stroke risk, and a strong correlation between plasma low-density lipoprotein cholesterol (LDL-C) and apolipoprotein B levels [26]. High HDL-C levels are linked to better prognosis [27]. Our study aligns with these findings, showing that high LDL-C, low HDL-C, and elevated TG levels are more likely to result in PSE. This can be understood as high cholesterol and triglyceride levels increase blood viscosity and contribute to vascular sclerosis, promoting clot formation [12][28][29]. Higher D-dimer levels indicate more significant brain tissue damage, increasing the likelihood of PSE. In general, lower activated partial thromboplastin time (APTT) and fibrinogen levels are associated with higher PSE risk, while INR, PT, and TT have a smaller impact. Among liver function indicators, aspartate aminotransferase (AST) had the greatest influence on PSE. High AST, low alanine aminotransferase (ALT), and low albumin levels also had some impact. Lingling Ding et al. found that liver enzyme subgroups defined by ALT and AST were linked to higher risks of adverse outcomes [30], which is consistent with our findings.

Studies have also shown that renal function biomarkers such as urinary microalbumin, cystatin C, and creatinine are associated with higher stroke recurrence rates and poorer prognosis [30]. In our study, low urea levels and high uric acid levels had a negative impact [31][32][33]. Our research supports these conclusions. Elevated uric acid levels at admission were positively associated with PSE, although patients with a prior diagnosis of hyperuricemia were less likely to develop epilepsy. Since uric acid acts as a strong antioxidant and has neuroprotective properties [34], patients with normal liver and kidney function and mild hyperuricemia may have greater resilience in emergencies [35][36]. However, excessively high uric acid levels suggest metabolic disorders and poor liver and kidney function, which are linked to a poor prognosis.

When stroke patients are admitted, cardiac enzyme tests are often conducted to rule out myocardial ischemia. However, studies have shown that elevated CK-MB in stroke patients may not be solely heart-related [37]. Cardiac enzymes are important prognostic indicators [38][39] and have been incorporated into stroke scores [40]. Some studies have reported a higher incidence of abnormal serum cardiac enzyme levels in the acute phase of stroke. While the abnormalities are not related to the stroke type, they are associated with stroke severity, with patients exhibiting consciousness disorders having a significantly higher incidence of abnormal cardiac enzymes than those without such disorders [41]. In our study, CK, CK-MB, and IMA in the cardiac enzyme profile had a significant impact and high predictive value, though further research is required to understand the specific mechanisms involved [34].

Although our study incorporated extensive clinical, imaging, and laboratory data to build more accurate prediction models using machine learning algorithms, surpassing traditional statistical methods, there were still several limitations in the modeling process.

While the current study offers valuable insights, the data sample may not be fully representative, and the model’s generalizability requires further evaluation. Although the data was collected from multiple tertiary hospitals and includes over 20,000 cases, earlier data was lost due to hospital system upgrades. The dataset mainly reflects patients diagnosed within the past five years and is predominantly from the Chongqing region, which may limit the model’s applicability to other geographic areas.

Additionally, the retrospective nature of the study led to the absence of some important predictive indicators. Many potentially valuable features, such as hemorheology, thromboelastography, and hormone levels, were missing and had to be excluded. Including these features could potentially improve the model’s accuracy.

To enhance the predictive power of the model, it would be beneficial to incorporate more data beyond baseline patient characteristics. The current analysis primarily used the results from the first examination upon admission, without fully utilizing information from subsequent exams. In future research, recurrent neural networks could be employed to extract features from the entire sequence of examinations more comprehensively.

To strengthen the study further, data standardization should be improved, and the number of cases and key indicators should continue to grow. Additionally, it would be advantageous to explore more advanced scientific methods, such as deep learning, and utilize all available data to improve prediction accuracy.

## Materials and methods

### Research patients

This study retrospectively included all stroke patients admitted to the Chongqing Emergency Center between June 2017 and June 2022 for the development of the prediction model. Data from three external validation centers—Qianjiang Central Hospital, Bishan District People’s Hospital, and Yubei District Traditional Chinese Medicine Hospital—were collected between July 2022 and July 2023 to validate and evaluate the model externally. The external validation cohort emphasized collecting positive cases to test the model’s ability to identify these cases accurately.

Inclusion criteria: (1) Age between 18 and 90 years at admission; (2) Diagnosed with acute ischemic stroke and hospitalized for treatment.

Exclusion criteria: (1) Patients with a history of stroke or transient ischemic attack (TIA); (2) Patients with a history of other conditions such as traumatic brain injury, intracranial tumors, or cerebral vascular malformations that may cause epilepsy; (3) Patients with a history of epilepsy or who have received antiseizure medications for the prevention of seizures or for other diseases (such as migraine or psychiatric disorders); (4) Patients who died within 72 hours after stroke onset.

This study collected de-identified data from relevant patients to build a multi-modal stroke patient database. The study protocol was approved by the Ethics Committees of Chongqing University Center Hospital, Chongqing University Qianjiang Central Hospital, Bishan District People’s Hospital, and Yubei District Traditional Chinese Medicine Hospital.

The selection process is outlined in Figure 1. A total of 42,079 records were retrieved from the stroke database, and 24,733 patients were diagnosed with ischemic or lacunar stroke with new onset. Hemorrhagic strokes (4,565), a history of stroke (2,154), TIA (3,570), unclear cause strokes (561), and records with missing essential data (6,496) were excluded. Patients whose seizures might have been caused by other factors (such as brain tumors, intracranial vascular malformations, or traumatic brain injury) (865), those with a seizure history (152), and patients who died in the hospital (1,444) were also excluded. Additionally, patients lost to follow-up (those without outpatient records or unreachable by phone) or who died within three months of the stroke incident (813) were excluded. Finally, 21,459 cases were included in the study.

**Figure 1.**
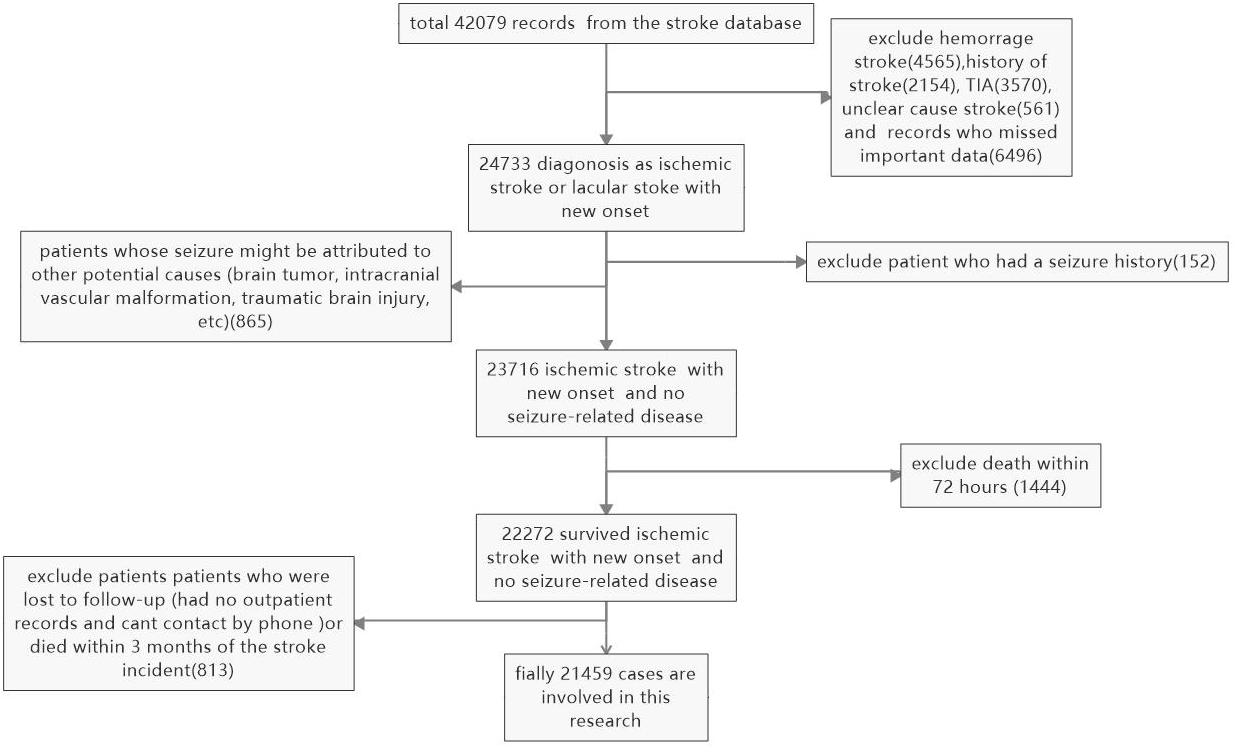
Selection and Exclusion Procedure of Patients A total of 42,079 records were retrieved from the stroke database, and 24,733 patients were diagnosed with ischemic or lacunar stroke with new onset. Hemorrhagic strokes (4,565), a history of stroke (2,154), TIA (3,570), unclear cause strokes (561), and records with missing essential data (6,496) were excluded. Patients whose seizures might have been caused by other factors (such as brain tumors, intracranial vascular malformations, or traumatic brain injury) (865), those with a seizure history (152), and patients who died in the hospital (1,444) were also excluded. Additionally, patients lost to follow-up (those without outpatient records or unreachable by phone) or who died within three months of the stroke incident (813) were excluded. Finally, 21,459 cases were included in the study.

**Figure 2.**
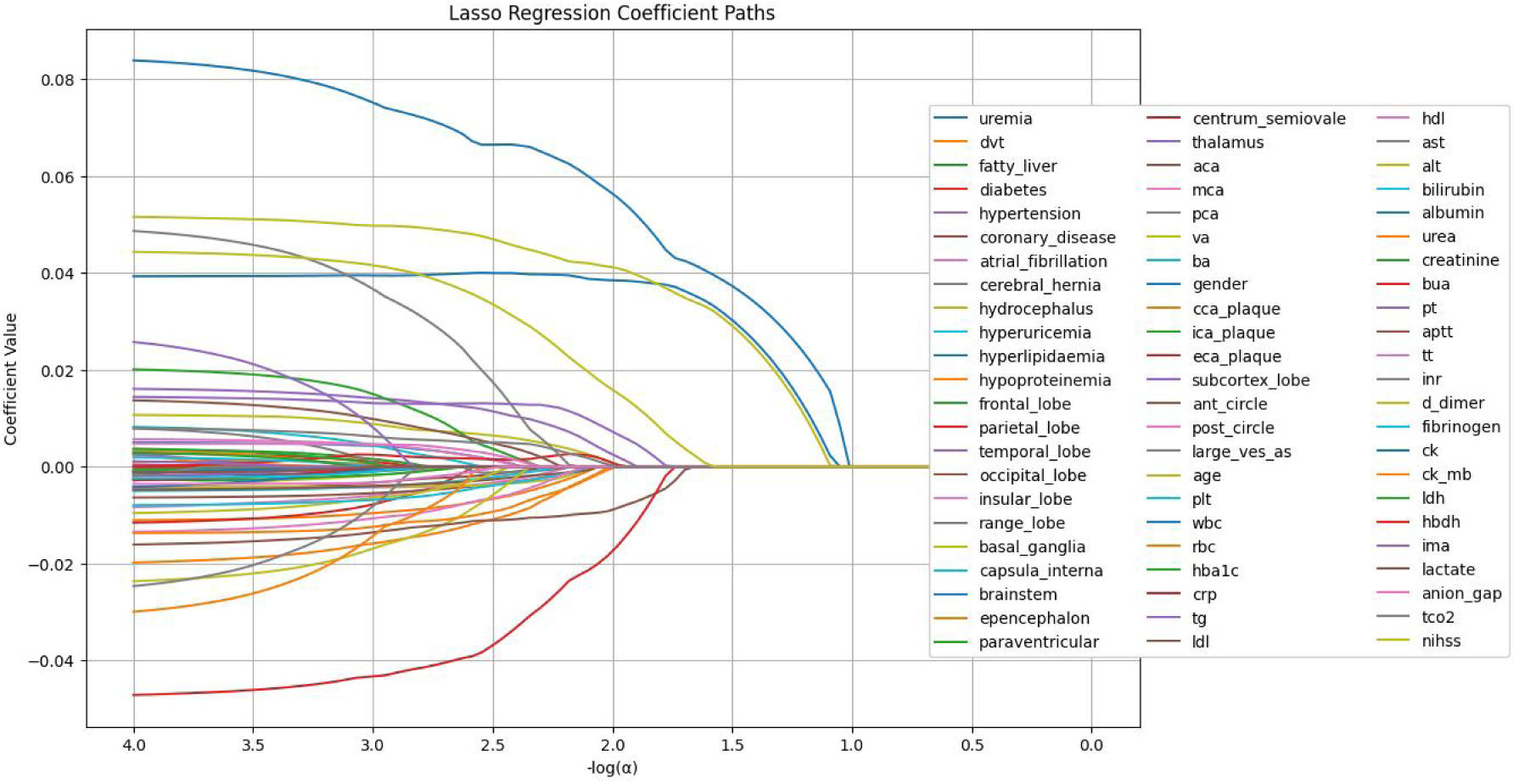
LASSO Regression Coefficient Paths The image shows the LASSO regression coefficient paths for various features related to a medical or research study. The x-axis represents the log of the regularization parameter alpha, and the y-axis shows the regression coefficient values. The lines in the plot represent the coefficient paths for different features as the regularization parameter changes. The features are labeled on the right side of the plot, and the most important features selected by the LASSO model are shown at the bottom of the image.

**Figure 3.**
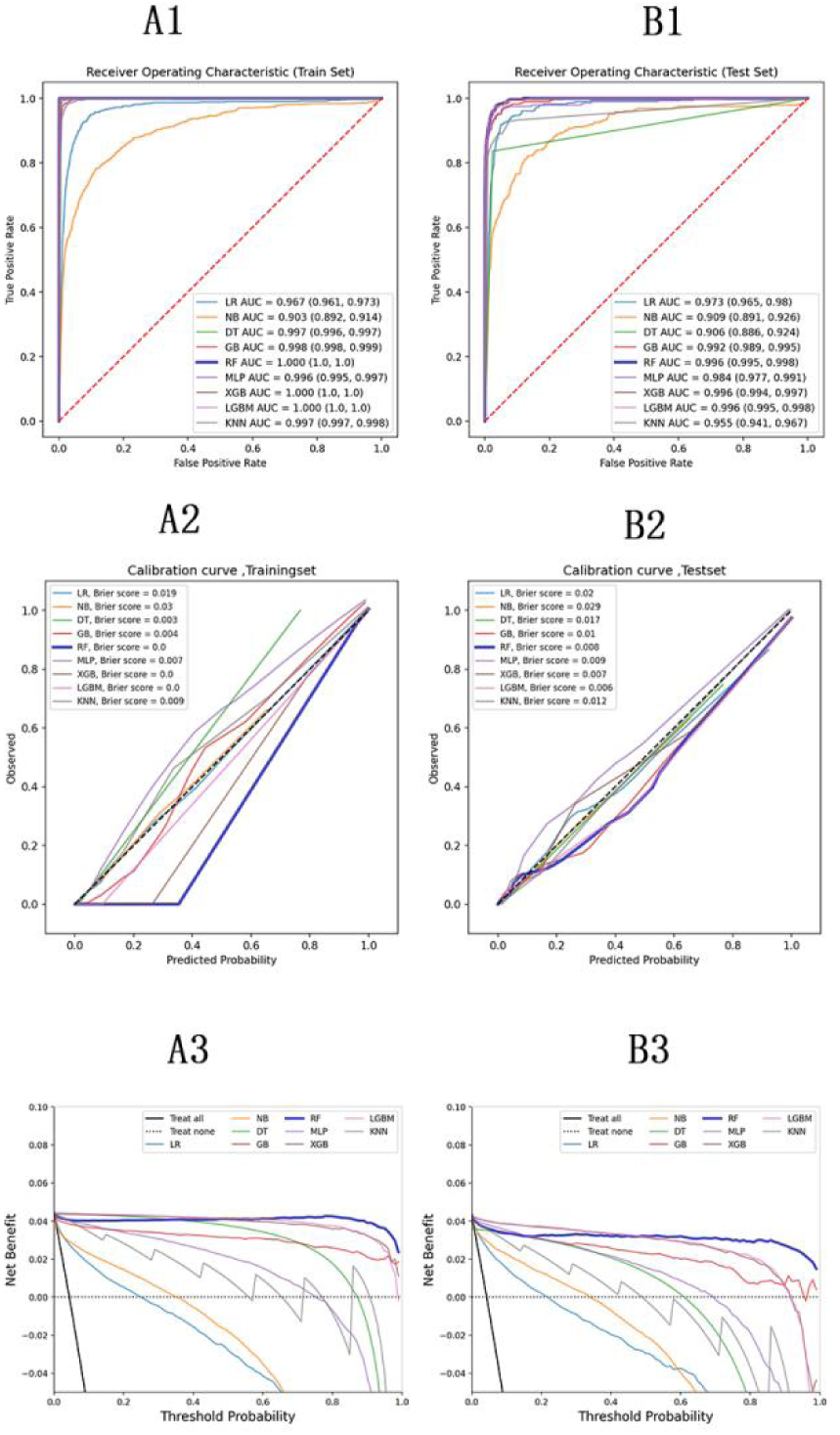
Model Evaluation Metrics and Curves The figure displays model performance curves across six sections (A1, A2, A3 on the left; B1, B2, B3 on the right) for training and test sets. ROC Curve: Illustrates the trade-off between sensitivity and specificity, with the AUC indicating overall model performance. Calibration Curve: Compares predicted probabilities to actual outcomes, assessing the model’s confidence accuracy. Precision-Recall Curve: Analyzes the balance between precision and recall at various thresholds, particularly useful for imbalanced datasets.

**Figure 4.**
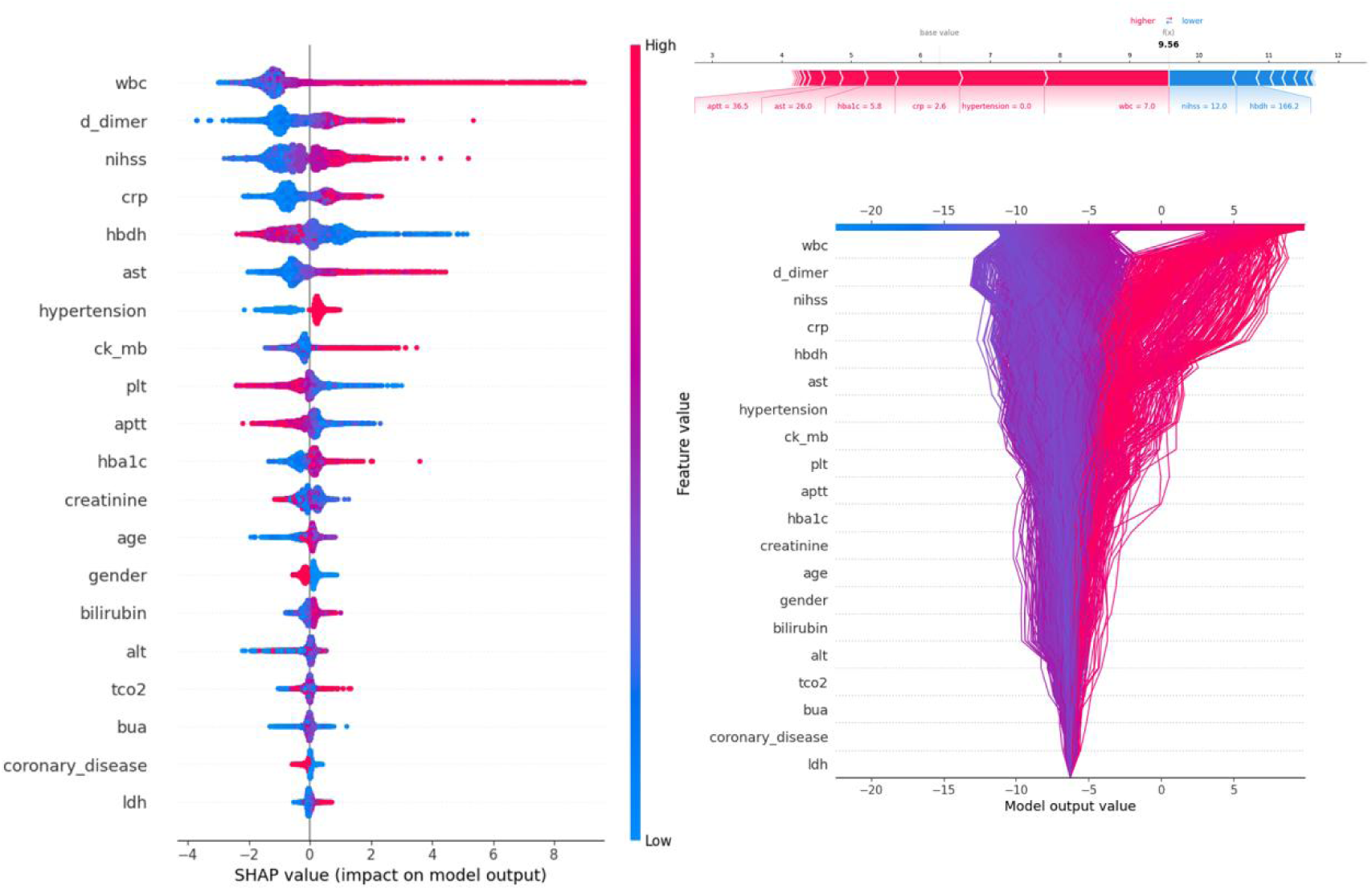
Description of the SHAP Values and Feature Importance SHAP Value (Left): Displays the impact of each feature on the model’s predictions, with features sorted by importance. The color gradient indicates the range of feature values, from low (blue) to high (red). Force Plot (Upper Right): Illustrates the contribution of individual features of the first sample to the final model output, highlighting how each feature value pushes the prediction away from the baseline value. Decision Plot (Lower Right): Visualizes the cumulative impact of features on the model output for each sample, showing how the feature values combine to produce the final prediction.

### Data collection

We extracted all relevant records and data from the hospital databases. Using PostgreSQL, we wrote Structured Query Language (SQL) to manage the data as follows:

1. General Information: This included gender, age, and NIH Stroke Scale (NIHSS) score at admission.
2. Comorbidities and Complications: These included uremia, previous deep vein thrombosis (DVT), diabetes mellitus, hypertension, coronary atherosclerosis, atrial fibrillation, cerebral hernia, hydrocephalus, hypoproteinemia, hyperuricemia, hyperlipidemia, internal carotid stenosis, and common carotid stenosis.
3. Brain Involvement (CT or MRI records): We recorded involvement of the cortical lobes and subcortical areas, including the frontal, parietal, temporal, occipital, and insular lobes, as well as the basal ganglia, internal capsule, brain stem, cerebellum, periventricular area, centrum semiovale, and thalamus. The extent of cortical involvement (frontal, parietal, temporal, occipital, and insular lobes) was scored, with each lobe contributing 1 point. Similarly, subcortical involvement (basal ganglia, internal capsule, brain stem, periventricular area, thalamus, and cerebellum) was scored with each area contributing 1 point.
4. Vascular Involvement (CTA, MRA, or DSA records): We recorded the presence of vascular stenosis or occlusion in the anterior cerebral artery (ACA), middle cerebral artery (MCA), posterior cerebral artery (PCA), vertebral artery (VA), and basilar artery (BA).
5. Key Laboratory Indicators: These included blood lipids such as triglycerides (TG), high-density lipoprotein cholesterol (HDL), and low-density lipoprotein cholesterol (LDL); liver function indicators such as alanine transaminase (ALT), aspartate aminotransferase (AST), bilirubin, and albumin; renal function markers such as urea, blood uric acid (BUA), and creatinine; blood gas parameters such as lactate, anion gap, and total carbon dioxide (TCO2); coagulation markers such as international normalized ratio (INR), prothrombin time (PT), activated partial thromboplastin time (APTT), thrombin time (TT), D-dimer, and fibrinogen; and myocardial enzymes such as creatine kinase (CK), creatine kinase isoenzyme (CK-MB), lactate dehydrogenase (LDH), ischemic modified albumin (IMA), and α-hydroxybutyrate dehydrogenase (HBDH).

### Data processing and model building

Processing of Missing Data: We recorded all laboratory indicators from the first set of tests after stroke admission (every stroke patient undergoes routine blood tests, and liver and kidney function assessments). Indicators with more than 10% missing data were excluded. The remaining indicators with missing values were imputed using the random forest algorithm with default parameters. We processed the features in order of missing values, starting with those that had the least missing data (as this requires the least information for imputation). When imputing a feature, missing values in other features were temporarily replaced with 0. After each regression prediction, the predicted value was inserted into the original feature matrix before proceeding to the next feature. Once all features were processed, the dataset was complete.

Distribution of Characteristics: We used univariate analysis to compare the distribution of characteristics between the PSE-negative and PSE-positive groups. The data were then divided into a training set and a test set in a 7:3 ratio.

Processing of Unbalanced Data: Given the low incidence of PSE and the small proportion of positive cases, we augmented the positive data in the training set using the Synthetic Minority Over-sampling Technique combined with Edited Nearest Neighbors (SMOTEENN). The SMOTEENN method from the imblearn Python package was applied with default parameters, and a random seed of 42 was set to ensure reproducibility.

Processing of Categorical Data: For categorical variables, we used the one-hot encoding method for transformation. We then applied the LASSO method to the training set to identify the most important features.

Model Building: First, we used LASSO regression to select the 20 most important features. We then employed 9 commonly used machine learning methods, including Naive Bayes, Logistic Regression, Decision Tree, Random Forest, Gradient Boosting, Multi-Layer Perceptron, XGBoost, LightGBM, and K-Nearest Neighbors. Hyperparameters for each model were optimized through grid search to enhance performance. Model evaluation metrics included accuracy, sensitivity, specificity, F1-score, positive predictive value, and negative predictive value. We also generated ROC curves, calibration curves, and decision curves to further assess model performance. An independent external validation dataset was used to evaluate the generalization ability of the selected model. Lastly, we applied the SHAP algorithm to interpret the best-performing model, analyzing the contribution of each feature to the model’s predictions and their clinical relevance. Through this process of model development, optimization, and interpretation, we constructed a machine learning model with strong predictive performance and interpretability, offering valuable support for clinical decision-making.

### Statistical approach

PostgreSQL v15 (http://www.postgresql.org/) was used to search and extract data from the local database. The open-source statistical package “Scipy.stats” in Python was used for statistical analysis. The details of the univariate significance analysis for each feature are as follows:

The Shapiro-Wilk test was applied to assess the normality of each feature’s distribution. For features that did not follow a normal distribution, the Mann-Whitney U test was used to evaluate their significance in relation to the target variable. For features with a normal distribution, the Levene test was performed to evaluate the homogeneity of variances. Features with homogeneous variances were analyzed using the Student’s t-test for significance, while those with heterogeneous variances were analyzed using Welch’s t-test.

Confidence intervals for AUC values and Brier scores were calculated using 1,000 bootstrap resampling iterations on the datasets. Binary classification thresholds for the predicted probabilities from all models were established using the maximum Youden index derived from the training cohort.

Throughout the study, a two-tailed p-value of less than 0.05 was considered statistically significant.

All the code used in this study was uploaded to https://github.com/conanan/lasso-ml.

## Conclusion

We developed an interpretable machine learning model to predict the risk of post-stroke epilepsy (PSE) in hospitalized patients with ischemic stroke. Using a large dataset of medical records, our artificial intelligence model demonstrates strong predictive performance for PSE. The key predictors identified by the model include NIHSS score, D-dimer levels, lactate levels, and white blood cell count, along with liver function and cardiac enzyme profile indicators. The model’s transparency and interpretability can build trust among clinicians and support decision-making. While the results are promising, further prospective studies are necessary to validate the clinical utility of this tool before it can be applied in real-world settings.

## Data availability statement

The codes,models,analysis results was uploaded at https://github.com/conanan/lasso-ml. The full dataset can be provided for researchers if needed by the corresponding author.

## Acknowledgements

The authors would like to thank the colleagues in the information and imaging departments for their hard work contributing to the final research results.

## Ethics approval statement

We confirm that we have read the Journal’s position on issues involved in ethical publication and affirm that this report is consistent with those guidelines.

## Patient consent statement

This study was a retrospective study and only deidentified patient data were collected, exempting the need for patient informed consent rights.

## Permission to reproduce material from other sources

There are no reproduce material from other sources.

## Clinical trial registration

The trail number is RS202406.

